# COVID-19 Outbreaks in Refugee Camps

**DOI:** 10.1101/2020.10.02.20204818

**Authors:** Carlos Hernandez-Suarez, Paolo Verme, Sergiy Radyakin, Efren-Murillo

## Abstract

We built a mathematical model for SARS-CoV-2 transmission and analyze it using both a deterministic and a stochastic approach. We used this model to project the burden of the disease in refugee camps characterized by peculiar demographic characteristics and a high level of deprivation, including lack of medical facilities and personnel, as well as limited possibility to implement containment and quarantine measures. Most of the parameters in our model were adapted from published literature but we used our own estimates of the basic reproduction number, *R*_0_ as well as the lethality by age group and gender. We projected the burden in terms of number of infections, number of deaths and number of bed-days in hospitalization and intensive care, among others. We conclude that the harsh conditions of refugee camps combined with a high share of young people leads to a relatively mild scenario for the burden of the disease.

## 2 Executive summary

### Objective

The report illustrates the results of a series of simulations of the spread of COVID-19 disease in refugee camps using a demo set of data from the Za’atari refugee camp in Jordan. The purpose of this exercise is to illustrate the type of information that can assist the UNHCR and camps’ administrators in preparing for a COVID-19 outbreak.

### Model

Simulations are conducted with a compartmental model that we call “SIZ”. This is a newly developed epidemiological model with eight compartments designed specifically to model COVID-19 outbreaks in closed and highly densely populated communities such as refugee camps. The model segregates those infected into symptomatic and asymptomatic categories and provides simulations using both deterministic and stochastic solutions. Whereas deterministic models are based on transition rates between compartments, stochastic models are based on transition probabilities between them. The main difference is that in a deterministic model an individual ‘splits’ among communicating stages, whereas in a stochastic model the individual moves to only one of those stages. Also, transitions from the current state **X**(*t*_0_) to the state **X**(*t*_0_ + *t*) are random, as well as the time for the transition, *t*. This allows for the estimation of standard deviations and confidence intervals. Solutions to the models have been programmed in Matlab, Python and Stata to check and validate results.

### Parameters

The basic reproductive number *R*_0_ used for these simulations is 3. This is a low-end reproductive number as compared to the literature on COVID-19 and a cross-country estimation prepared by the authors based on publicly available information. The rest of the parameters in the model such as residence time in each compartment and transition probabilities between compartments were estimated directly or indirectly from the existing literature or using information collected from a data set from Mexico [10] which has been used to benchmark lethality rates by age and gender.

### Policies

Some plausible control policies were included in the model including use of masks, contact tracing and quarantine, promoting social distance and reducing infection time by self-awareness. Results are presented for four sets of parameters described in the Results sections. Two of the simulations include contact tracing policies and two exclude them, and for each of these options we simulate the baseline contact rate (no reduction in contact rate) and a 33% reduction in contact rate, which can be the result of different distancing or isolation policies. Pharmaceutical treatments are not included in the model since there is no known-effective therapy. Nevertheless, if one wishes to do so, the parameters *p* and *w* in the model can be changed to reduce the lethality of the disease. The same applies for vaccines which would require a reduction in the contact rate.

### Results of the Deterministic Solution

Results of the deterministic solutions are shown in tables 12 to 15. It is shown that, with no reduction in contact rate and no contact tracing, 95% of the population will be infected, although the mortality will be comparatively low (less than 2 in 1000) mainly because of the large share of young people among refugees (60 % is less than 20 years old in the Za’atari refugee camp). The simulations also show that 3/4 of the infections will be reached in about 10 weeks. Since the time to first death is also about 7 weeks, the epidemic will not be noticeable for the first month and a half and will be running underground, with an estimate of 25 % of infections by week 8.

Most of the disease will manifest itself in hospitals and intensive care units. The simulations reveal that, in the worst case scenario of those studied, 1,806 and 1,052 bed-days^1^ are required in hospital and Intensive Care (IC) units respectively. With the average time of hospitalization used (three days), 49 beds in hospitals and and 25 in IC units will be required to fulfill most of the demand.

If we perform contact tracing and quarantine 1*/*3 of the infected individuals, the epidemic size will drop from 94% of the population to 85%. If we reduce the contact rate by 1*/*3, the epidemic size is reduced to 79% of the population. Both strategies combined will reduce the epidemic size to 58%, although achieving this reduction would be difficult in refugee camps [23].

The analysis suggests that it is important to concentrate efforts on reducing the effective contact rate by encouraging the use of face masks and promoting social distance, whereas testing and contact tracing is a less effective strategy due mainly to the high *R*_0_ value. Also, promoting a reduction in the infectious time through education and self-awareness is of little effect, mainly because the fraction of symptomatic individuals requiring hospitalization is small, and the epidemics is clearly driven by those asymptomatic or not requiring hospitalization.

Lock-downs are effective to avoid the virus from entering the camp. But the likelihood that this can be actually implemented in an effective way is dubious. The disease will enter the camp if there is some kind of interaction between the camp and any city or village nearby where an outbreak has began. Since a small fraction of individuals will develop symptoms that require hospitalization, this means that, on average, there will be many infections before the first infected individual is noticed (the time to first death is on average over 6-7 weeks), which makes a lock-down ineffective. Individual isolation is also difficult [23], mainly because it requires the isolation of at least all members of a family.

### Results of the Stochastic Solution

The average number of deaths and bed-days calculated in the stochastic model are similar to those obtained for the deterministic model.^2^ Nevertheless, there are important differences in the maximum number of hospitalized in a day as well as in the number of beds required to cover the demand of hospital and ICU services. Other differences are related to the time to some events such as the time to reach a given percentage of infections. In a stochastic model, we are discarding those simulations in which the epidemic vanished quickly with no deaths, thus, the distributions are switched to the right of the deterministic model and the time to the occurrence of an event is larger.

In the results sections, we also present figures illustrating the number of beds required to fulfill the demand for bed-days, also including the margins of error. For example, we can see from Figure 7 (top panel) that, with 83 beds, it would be possible to satisfy 90% of the demand for hospital beds, whereas 48 beds would be required to satisfy 90% of the demand for beds in Intensive Care Units (ICUs, bottom panel). Standard errors are included in all figures and represented by vertical bars.

### The Web Application

The stochastic solution of the SIZ model is also available on the web where it is possible to reproduce the results presented in this study, or run additional simulations by varying the parameters of interest. The web app is currently hosted at this address:

https://bit.ly/3jdtOXX

### Validation of results

In an attempt to benchmark and validate our results, we compare them with one other similar study conducted in a refugee camp in the Cox’s Bazar district of Bangladesh, and with the information emerging on COVID-19 outbreaks in refugee camps.

In our knowledge, there is only one other study that estimated the potential impact of COVID-19 in refugee camps. This is a study conducted by John Hopkins University on the Kutupalong-Balukhali Extension Site, Cox’s Bazar, Bangladesh [23]. We first compare the results of this study with the results of our study on the Za’atari refugee camp in Jordan and then compare the two models used by these studies applying our SIZ model to Kutupalong-Balukhali Extension Site. To note that the JHU-SEIR model also estimates the probability of an outbreak (with probabilities ranging from 61 to 92%), whereas the WB-SIZ model only considers outcomes following an outbreak.

Table 1 reports the key parameters and results of the two studies. The two studies result in very similar estimates on the population shares of infected people at the end of the epidemic. The JHU study for the Cox’s Bazar refugee camp reports 91.1% infection rate with an *R*_0_ comprised between 2 and 3 whereas our study reports an infection rate of 94.2% with an *R*_0_ of 3. Similarly, our model applied to the Kutupalong-Balukhali Extension Site finds an infection rate of 94.3%. Essentially, all simulations concord in finding that, at the end of the epidemics and with no pre-existing immunity (something that is still debated in the literature), the quasi-totality of people are expected to be infected.

**Table 1:**
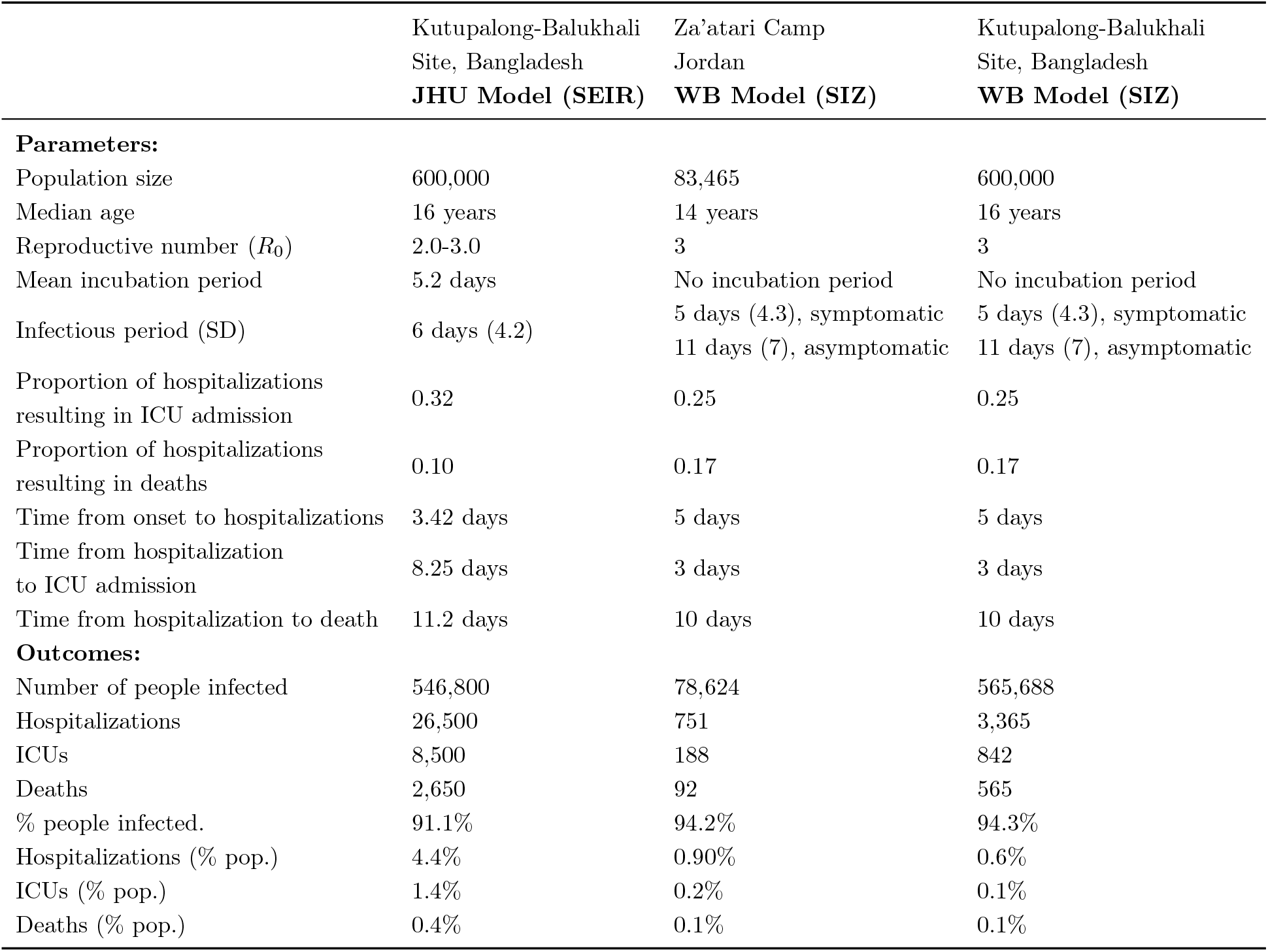
Parameters and outputs comparison between JHU SEIR model [23] and WB SIZ model.

**Table 2:**
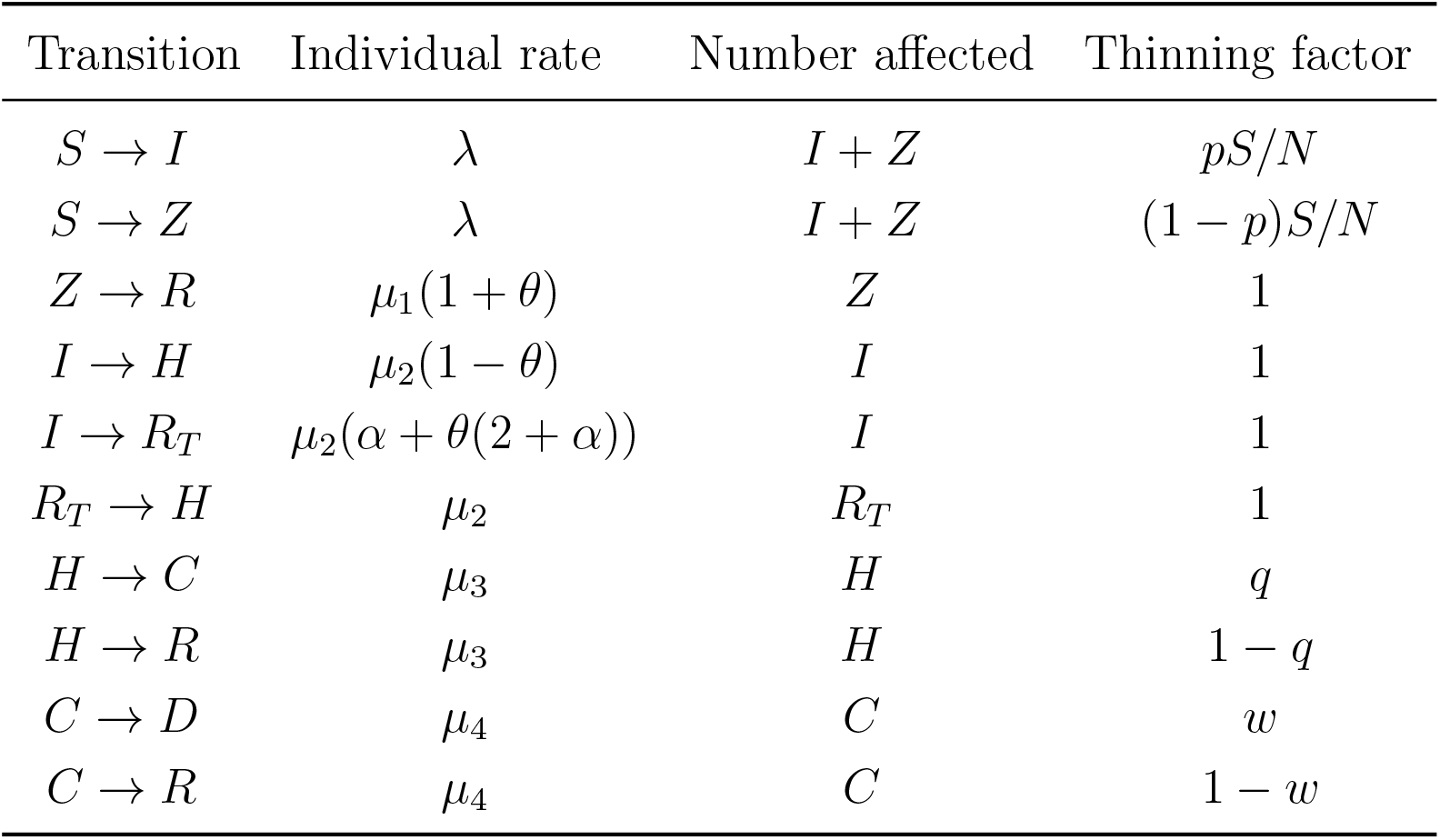
Transition rates

However, there are significant differences across the two models in terms of overall numbers of hospitalizations, hospitalizations in Intensive Care Units (ICUs) and deaths. Both studies find relatively small shares of people in need of hospitalization (between 0.6 and 4.4 %), ICUs (between 0.1 and 1.4%), and low death rates (between 0.1 and 0.4 %), which are rates consistent with the latest literature. It is also understood that these low rates are mainly explained by the relatively young age of the population living in refugee camps. However, the the JHU study provides estimates that are much higher than our estimates. The main explanations for this difference are the benchmark data used by the two studies for the transitions between infections, hospitalizations and death and the age adjustments. Our study relies on a comprehensive data set of individual observations from Mexico, whereas the JHU study relies on information available in the literature. As it is uncertain what best applies to refugee camps, the most suitable interpretation of these differences is in terms of ranges of possible outcomes. This implies that preparations for the outbreak should rely on the upper estimates.

It is also possible to compare results of these simulations with the observed evolution of the spread of the disease in the Za’atari refugee camp in Jordan and the Cox Bazaar refugee camp in Bangladesh. According to the refugee response coordination platform in Jordan, testing in the Za’atari refugee camp has been underway and a facility for treatment of COVID-19 cases has been already built in the expectation of an outbreak. However, as of July 13th, 2020, no positive cases were detected^3^ Therefore, either the virus has not yet entered the camp or it is still in an very early stage when casual testing is unlikely to catch positive cases. In either case, the situation on the field cannot provide any information to validate the simulations proposed in this study.

Some initial comparisons can instead be done between the results of the [23] Kutupalong-Balukhali Site simulations and the evolving situation on the ground. The first refugee COVID-19 case in the Cox’s Bazaar refugee camp was registered around May 15th, 2020^4^, and, according to the WHO “*As on 28 June, as many as 2,456 cases of infection have been confirmed in Cox’s Bazar District and 50 refugees have tested positive in the Rohingya camps*.”^5^ Therefore, the virus is widely present in the district and in the camp, particularly if one considers the still limited number of tests per day conducted in the region and the number of observed infections went from 1 on May 15th to 50 on June 28th.

We know from studies worldwide that the true number of infections may be as large as ten times what is observed, even in countries where testing is widespread. According to the simulations provided by [23], “*On average, in the first 30 days of the outbreak, we expect 18 (95% prediction interval [PI], 265), 54 (95% PI, 3223), and 370 (95% PI, 41,850) people infected in the low, moderate, and high transmission scenarios, respectively*.” The study focused on the Kutupalong-Balukhali Expansion Site which is home of about 600,000 of the more than 1 m refugees in the Cox’s bazaar district of Bangladesh, and we do not know how many of the 50 infected cases were found in this particular zone. However, with a rough estimate of 30-40 cases, it is entirely plausible that in the 43 days that passed between May 15th and June 28th, around 300-400 people have been infected in the camp. This would be within the range found in [23]. The WB-SIZ model applied to the Kutupalong-Balukhali Expansion Site results in similar infection rates at the end of the epidemic but reaches the end much faster than the JHU-SEIR model. This means that we would have expected the spread of the infection to be more advanced by day 43. This may depend of course on the effectiveness of the measures taken in the camp but, regardless of these measures, we should expect continued growth of infections in the next few months.

These findings warn against complacency with the low numbers of COVID-19 infected persons currently observed in Jordan and Bangladesh. Once the virus enters the camp, the simulations in [23] and our study show that the virus can spread quickly and widely, potentially reaching over 90% of the population. Thanks to the age and gender profile of the camps, the number of hospitalizations and deaths will be low relatively to older populations, but the numbers may still overwhelm hospitals and health care facilities. In this respect, preventive measures such as building COVID-19 field hospitals as in the Za’atari refugee camp should be supported and not regarded as excessive measures.

## 3 The SIZ model

### 3.1 Compartments

The model proposed is an epidemiological compartimental model structured into 8 compartments as illustrated in Figure 1.

**Figure 1:**
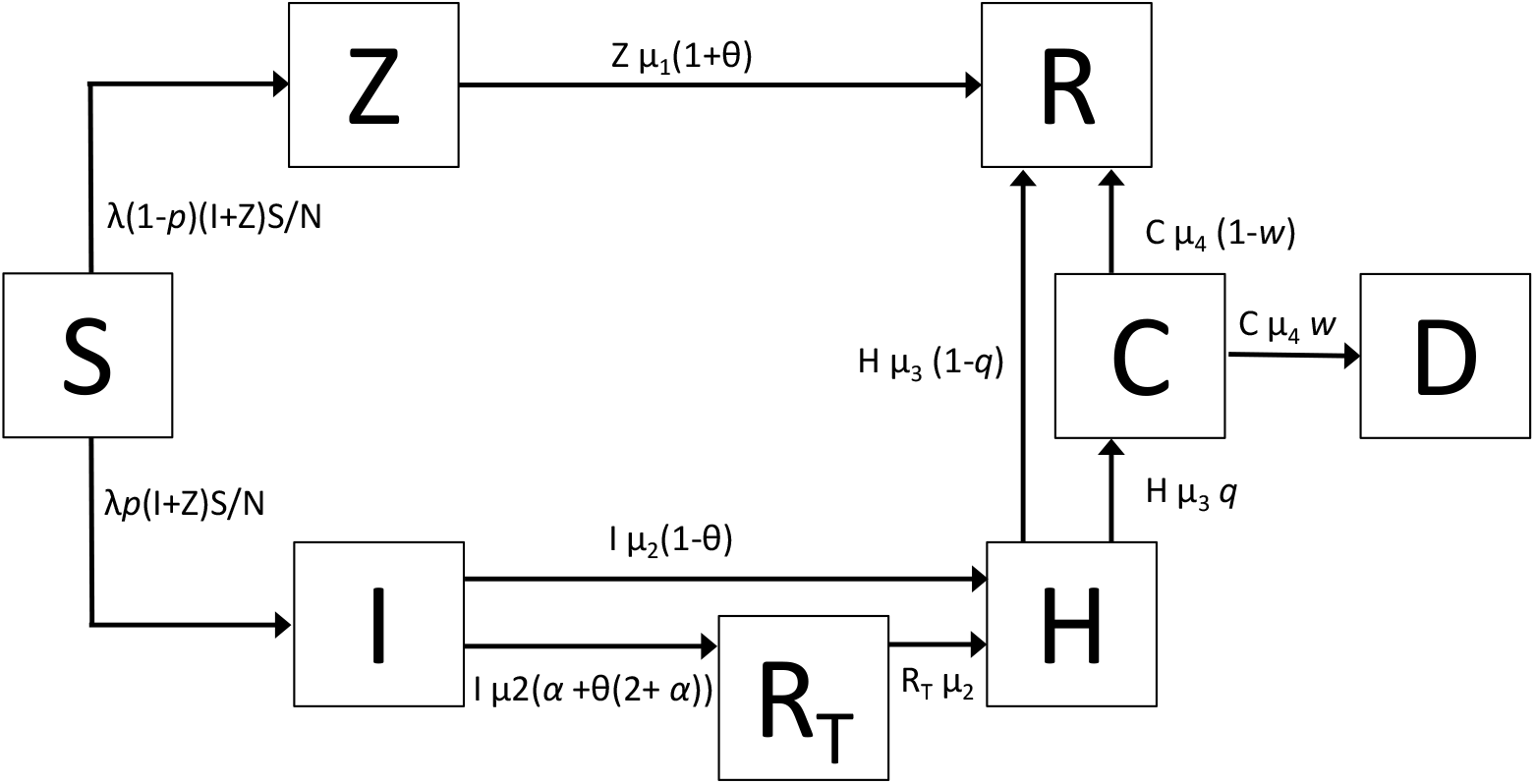
The epidemic model adapted. *S*= susceptible; *I* = symptomatic infected that will need hospitalization eventually; *Z* = asymptomatic infected that will not require hospitalization; *H*=hospitalized not in intensive care; *C*=intensive care; *R* = recovered; *R*_*T*_ = removed temporarily; *D*=dead. *N* = *S* +*Z* +*I* +*H* +*R*+*R*_*T*_ +*C* +*D*.

**Figure 2:**
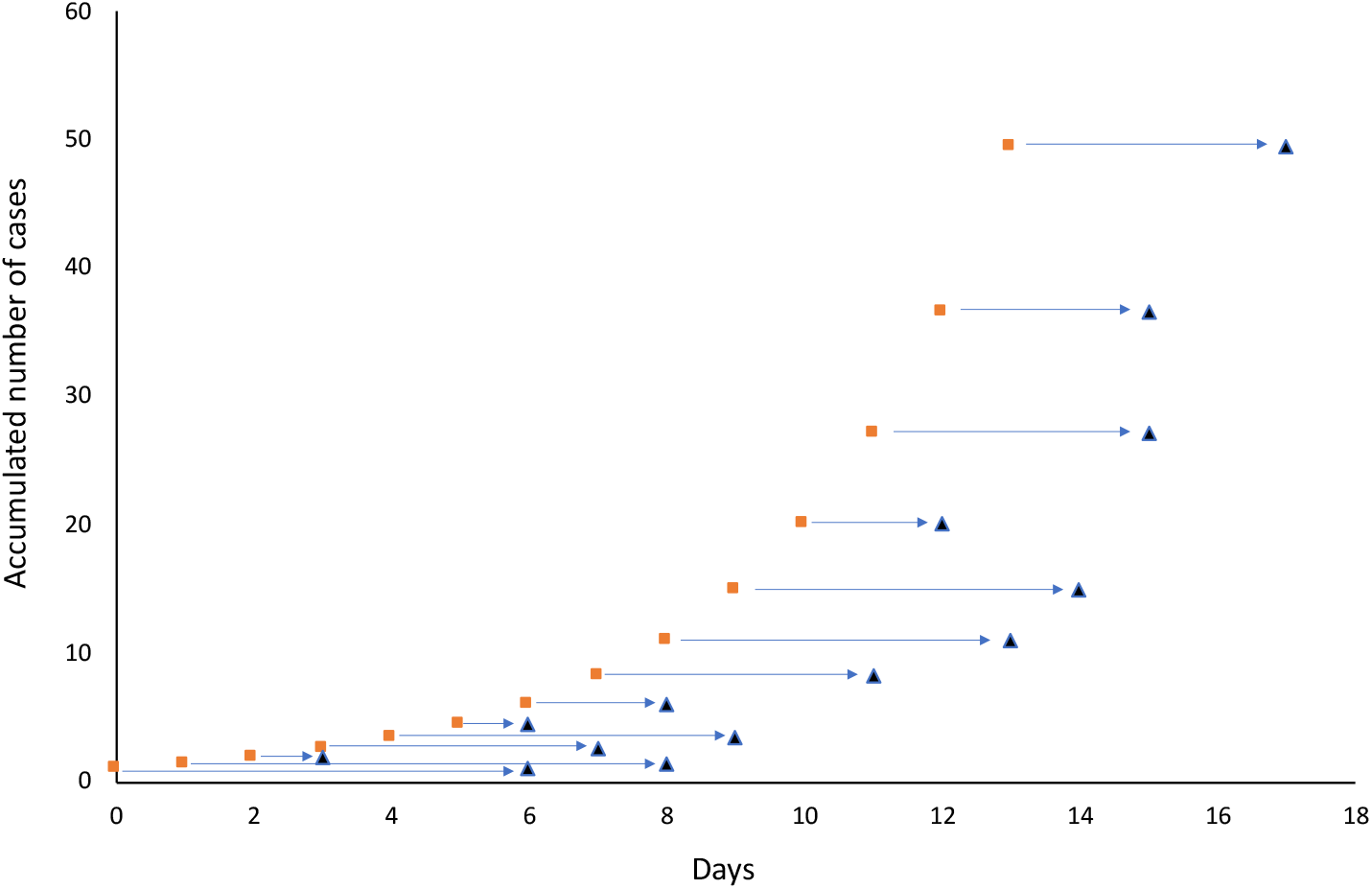
An illustration of the difference between time of infection: real (squares) and observed (triangles). The time of confirmation depends on many factors among others: availability of tests, symptom onset, having documented contacts with confirmed cases, etc.

The compartments are described as follows:

1. All individuals start as susceptible, *S*. Once an individual is infected, it becomes infectious immediately, either in category *I* (symptomatic) or *Z* (asymptomatic). By symptomatic we define those individuals that will develop eventually severe symptoms that will require hospitalization, whereas an asymptomatic individual refers to those that will develop none or mild symptoms that will not require hospitalization. The probability that an infected starts as symptomatic is *p* while it starts as an asymptomatic with probability (1 *– p*).
2. Infected asymptomatic *Z*, stay infected and infectious for an average amount of time 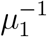. When they leave the *Z* stage they move immediately to stage *R* (removed) where they are alive, non-infected and non-infectious. Individuals in stage *R* are assumed to be immune, and they play no role in the dynamics of the epidemic. An individual in stage *Z* can also be moved to stage *R* without finishing the infectious stage if tested positive for SARS-Cov-2 virus. We assume that testing is implemented according to a *cordon sanitaire* where some contacts are tested for SARS-Cov-2. Testing depends on a parameter *θ* that will be described later.
3. Infected symptomatic *I* stay infected and infectious for some time and will develop eventually severe symptoms that will require hospitalization. An individual in stage *I* will move out of the *I* stage for two reasons: either this individual developed severe symptoms and moves to hospitalization *H* which occurs at a rate *µ*_2_ or it moves to temporary quarantine *R*_*T*_ which occurs at a rate *µ*_2_*α*. Parameter *α* reflects the possible effect on self-awareness. When individuals are more aware of the epidemic, its more likely that they will attribute classical symptoms of the disease as being infected, thus may reduce contact rate and self-isolate. *α* = 0 implies individuals keep infectious until they develop severe symptoms and move to hospitalization. *α >* 0 implies individuals will stop being infectious by self-awareness and self-isolation before the onset of severe symptoms.
4. Infected symptomatic *I* may also leave stage *I* by testing, as it was described for individuals in stage *Z*. The only difference is that individuals tested positive in stage *I* do not move to stage *R* since they are quarantined in a compartment *R*_*T*_ temporarily, where they remain until the appearance of severe symptoms that will require hospitalization. Testing depends on a parameter *θ* that will be described later.
5. Individuals removed temporarily in *R*_*T*_, move to hospitalization *H* at a rate *µ*_2_, due to the memoryless property of exponential distributions.
6. Hospitalized individuals *H* remain hospitalized for an average amount of time 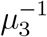 and they will need intensive care *C* with probability *q* or recover *R* and become immune with probability 1 *– q*.
7. Individuals in intensive care *C* stay in this stage for an average time 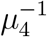 and may die with probability *w* or recover and become immune with probability (1 *– w*).

### 3.2 Parameters

The parameters in the model are:

- *λ*, the contact rate, that is, the average number of contacts per unit of time.
- 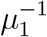, the average time in the compartment *Z*.
- 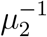, the average time in the compartment *I*.
- 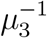, the average time in hospitalization (not in intensive care).
- 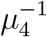, the average time in the intensive care unit.
- *p*, the probability that an infected individual becomes a symptomatic.
- *q*, the proportion of hospitalized individuals that will require intensive care.
- *w*, the proportion of individuals in intensive care that die from the infection.
- *θ*, the fraction of infected contacts that are removed due to testing. *θ* = 0 implies no testing, while *θ* = 1*/*2 half of the infected contacts of an individual that has been tested positive, are removed.
- *α*, is the effect on the exit rate from compartment *I* by symptom awareness and self isolation. *α* = 0 means no reduction in average in stage *I* due to self-awareness and isolation. The average time in stage *I* (excluding testing effects) changes from *µ*_2_ to *µ*_2_(1 + *α*).

### 3.3 Comments on the model and parameters

- The model does not consider infections caused by individuals in stages *H* and *C* (Hospitalized). This has been documented, but nosocomial infection is reduced in comparison to the rest of the susceptible.
- The model assumes implicitly that there is never shortage of beds in hospitals, including intensive care units. It is, therefore, an approximation.
- In the model, stages *I* and *Z* are separated for three reasons:
  - Individuals in stage *I* can reduce the average time in that stage with appropriate information, tending to be alert to the onset of symptoms which will allow them to self-isolate totally or partially. This is controlled by *α*.
  - It is important to keep the ratio *Z/*(*I* + *Z*) = 1 *– p*, that is, we need to observe the fact that a proportion of those infected are asymptomatic.
  - Separating *I* and *Z* would allow to change the infectiousness of the two kind of infected, if necessary.
- Although it is clear that the outcome of the epidemics is affected by the age structure of the population (seems that lethality is higher in older people), if we are willing to concede that all age categories are equally likely to become infected [24] but they respond differentially to the disease, then there is no need to write a model that include a compartment for each age category (with the necessary associated parameters) but to apply the distribution of differential lethality *a posteriori*. This will be explained in detail later in this report.
- The model lacks of a *latent* or *exposed* class for two reasons:
  - There are reports [20] that infected individuals become infectious quickly, sometimes in the first 1-2 days.
  - A latent class does not change *R*_0_ nor the final epidemic size, only delay events in the rest of the compartments a little bit.
- It is clear that the model cannot be solved explicitly, in the sense that we cannot derive analytic expressions for *S*(*t*), *Z*(*t*), *I*(*t*), *H*(*t*), *C*(*t*), *R*(*t*), *R*_*T*_ (*t*) and *D*(*t*). We will solve those expressions numerically and simulate a stochastic version of the model. But we will find analytic expressions for the long range behaviour of *R*(*t*) and *D*(*t*).
- All parameters above will be estimated or deducted from medical literature so far, except *λ*.
- Testing in the model refers to testing for actual infection with SARS-CoV-2, not for IgG or IgM (immunity tests). Laboratory testing always is implemented several weeks after the outbreak when the strains are identified and kits produced, so, it is necessary to set *θ* = 0 at the beginning of the epidemic.
- Parameter *θ*, the testing capacity needs special explanation: it is assumed that when individuals in stage *I* develop symptoms, they are tested and some of its contacts are tested. *θ* is the fraction of all infected contacts that are removed from circulation by testing. This shortens the infectious time of tested individuals (independently if they are in stage *I* or *Z*) by half, by property of exponential distributions.
- Choosing an appropriate parameter *α* is as follows: the average time spent in stage *I* with no testing is *µ*_2_ + *αµ*_2_ = *µ*_2_(1 + *α*). If we assume that certain informative policy may reduce the average time to a fraction *f* of its original value, then *α* = 1*/f –* 1.

## 4 The Deterministic Solution of the Model

### 4.1 Transitions between compartments

All rates between compartments are the product of three elements: an individual event rate, the number of individuals affected by the previous rate and a *thinning* probability.

Since the individual rate is the rate of a Poisson process, the resulting product is the rate of a Poisson event which is important for the future stochastic simulations. In this model, it is assumed that both types of infected, *I* and *Z* are equally infectious.

### 4.2 The differential equations

The set of differential equations for the model in Figure 1 (eliminated the time index for simplicity) is:

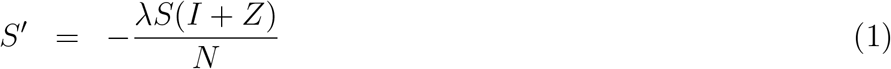

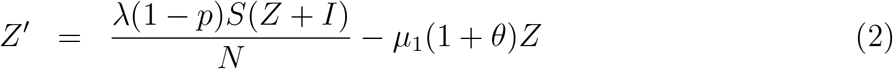

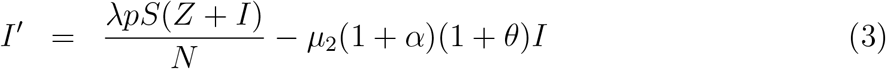

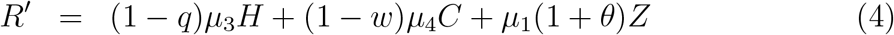

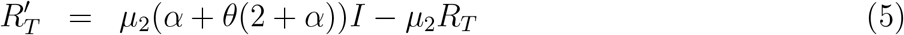

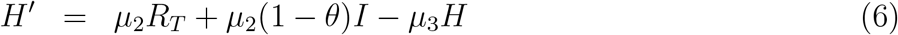

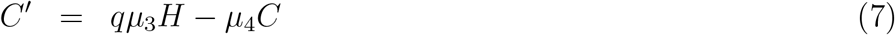

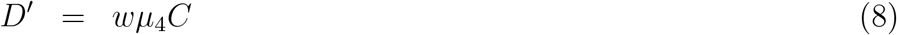

with initial conditions *S*(0) = *N –* 1, *Z*(0) = 1, all others equal to 0.

### 4.3 Adjusted lethality by age group and gender

Parameter *p* in the model, the probability that an individual will eventually develop severe symptoms that will require medical attention, is one of the most important parameters that define the outcome of the epidemic in terms of fatalities. The relevance of *p* is that it is a property of the disease. However, *p* seems to depend on the age of the infected, *p* being higher for older individuals. In this section, we calculate the lethality for age group which will allow us to estimate a global *p* at the population level. We will then show how to obtain the *p* for a population with a different age structure, as in refugee camps.

Tables 3 and 4 show the characteristics of hospitalization, intensive care and deaths by age group for Mexico’s COVID-19 epidemic using Mexico’s IMSS database^6^. Column **f** contains the relative frequencies of each age group. Column **h** shows how the hospitalizations where distributed among the different age groups. Column **c** shows, for each age group, the proportion of individuals that needed intensive care from those who went to hospital. Column **d** shows, for each age group, the proportion of individuals that died from those who went to hospital.

**Table 3:** Male demographics (**f**), hospitalization (**h**), intensive care (**c**) and death (**d**) rates, raw lethality (**u**) and adjusted lethality (***θ***)

| Males |  |  |  |  |  |  |
| --- | --- | --- | --- | --- | --- | --- |
| Age | **f** | **h** | **c** | **d** | **u** | **$\theta$** |
| 0-9 | 0.098713 | 0.004506 | 0.000000 | 0.185185 | 0.000834 | 0.000028 |
| 10-19 | 0.099764 | 0.003004 | 0.000000 | 0.166667 | 0.000501 | 0.000017 |
| 20-29 | 0.081298 | 0.093625 | 0.005941 | 0.049911 | 0.004673 | 0.000155 |
| 30-39 | 0.072028 | 0.204940 | 0.003988 | 0.119707 | 0.024533 | 0.000811 |
| 40-49 | 0.055658 | 0.226469 | 0.006434 | 0.266765 | 0.060414 | 0.001998 |
| 50-59 | 0.038507 | 0.218959 | 0.002611 | 0.489329 | 0.107143 | 0.003543 |
| 60 + | 0.042181 | 0.248498 | 0.018201 | 0.714574 | 0.177570 | 0.005871 |

**Table 4:**
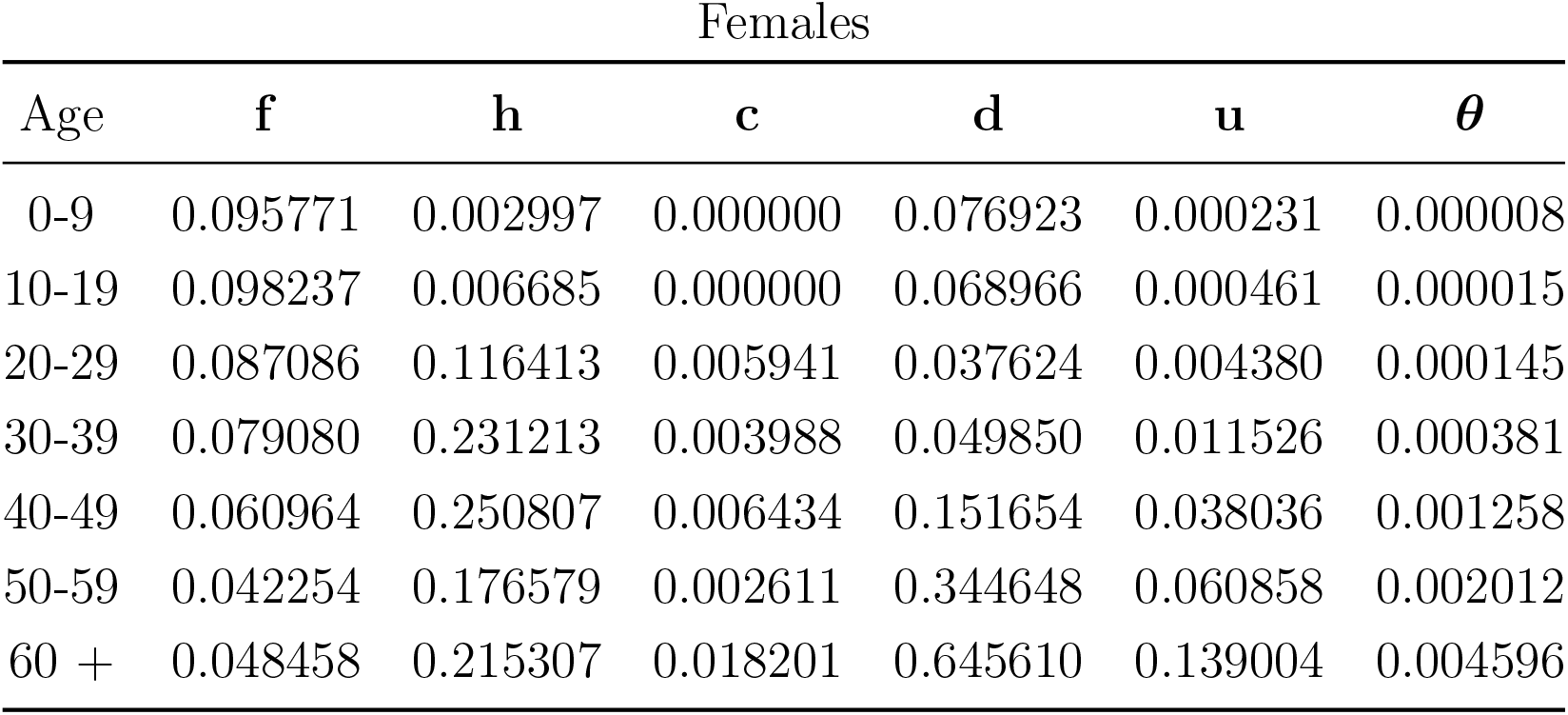
Female demographics (**f**), hospitalization (**h**), intensive care (**c**) and death (**d**) rates, raw lethality (**u**) and adjusted lethality (***θ***)

From these, we obtain the *raw lethality* by age group **u** = **h** *º* **d**. Overall lethality is **f**’_1_**u**. Nevertheless, we cannot estimate the lethality with **f’** _**u**_ since we are including only data from individuals that went to the *I* compartment and then to hospitalization whereas lethality should consider all infections. In other words, we do not have information on how many people went to compartment *Z*.

Estimates of the overall lethality are in the range 0.001 *–* 0.003 [21, 8]. We will use the lethality reported by [8] of 86 in 100, 000 (about 0.001) because it is a more recent study with a large number of blood donors (the possibility that conditions in refugee camps can worsen the severity of the outbreak will be considered later in our model).

Based on this lethality, we need to adjust **u** so that **f** ^*/*^**u** = 0.001, which implies a normalization factor *k* = 0.001*/*(**f** ^*/*^**u**) = 0.0347. We then obtain the *adjusted lethality* ***θ*** as ***θ*** = *k***u**. By performing this analysis by gender and age group, we obtain Tables 3 and 4. The interpretation of adjusted lethality is as follows: if we select a male person at random from the population in age group 30 *–* 39, the probability that this person will die from COVID-19 given that the person is infected is 0.000811, whereas if it is a female, the probability is 0.000381

#### 4.3.1 Adjusted lethality in the Za’atari refugee camp

If the information necessary for the estimation of the adjusted lethality rate is not available, as for areas not yet reached by the virus such as the Za’atari refugee camp, one can use the adjusted lethality of a reference population. For example and for the case of Za’atari camp, we can use the adjusted lethality estimated for Mexico by age group and adjust these lethality rates using the age and gender structure of the Syrian population living in the camp. The composition of the population structure of Syrian refugees in Jordan is shown in Table 5. Using this age and gender structure, the adjusted lethality for the camp is obtained with:

**Table 5:** Population structure of Syrian refugees in Jordan

| Age | $f_M$ | $f_F$ |
| --- | --- | --- |
| 0-9 | 0.1934 | 0.1823 |
| 10-19 | 0.1190 | 0.1116 |
| 20-29 | 0.0714 | 0.0794 |
| 30-39 | 0.0574 | 0.0611 |
| 40-49 | 0.0337 | 0.0348 |
| 50-59 | 0.0127 | 0.0171 |
| 60 + | 0.0110 | 0.0150 |
| Total | 0.4987 | 0.5013 |

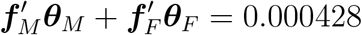

which is about 2.3 times smaller than the calculated from the Mexican IMSS data, due to the higher relative frequency of younger ages.

### 4.4 The parameters *p, q* and *w*

The adjusted lethality that we estimated for the Za’atari camp (0.000428) starting from the IMSS data can be expressed as the product *pqw*. From the same IMSS data, we can also estimate the product *qw* (the fraction of individuals that will die, given that they entered the *I* compartment) as:

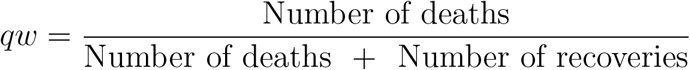

which is 0.1675 (note that the number of recoveries is only among those that went through the *I* compartment). It is then straighforward to estimate *p* as *p* = *pqw/qw* = 0.000428*/*0.1675 = 0.0026. This implies about 1 in 400 infected will become symptomatic infected, in this particular camp. The fraction of hospitalized that require intensive care (*q*) is derived from data on Iceland^7^, where this parameter is estimated at about 0.25. Finally, from *q*, we can derive *w* as *qw/q* = 0.1675*/*0.25 = 0.67.

### 4.5 Estimation of the exit rates *µ*_*i*_

The exit rates are the inverse of the average time in a given stage. For the deterministic model, if *L* is the average time in some state, then the exit rate *µ* = 1*/L*.

#### 4.5.1 Infected symptomatic (I)

The most recent and comprehensive report on the duration of the period a person remains infectious comes from the National Centre for Infectious Diseases in Singapore^8^. This study suggests that this period may begin around 2 days before the onset of symptoms, and persists for about 7 - 10 days after the onset of symptoms. That is a total period of between 9 and 12 days. However, people who are symptomatic become aware of the infection by definition and should be expected to take measures such as isolation to contain the spread of he virus. If this factor is taken into account, one can estimate the period of infectiousness at 5 days as suggested in [13]. See also [2] and [15]. Therefore, we take *µ*_2_ = 1*/*5

There is no data on the variance of this parameter, but if we use a gamma distribution and consider that subjects have no more symptoms after 12 days (see [13]) we can estimate a standard deviation. We need *n* and *γ* such that *n/γ* = 5 and *CDF*_*X*_(*n, γ*; 12) = 0.95. This yields *n* = 6, *γ* = 1.1765, for an average time of 5 days and a standard deviation of 4.33.

#### 4.5.2 Infected asymptomatic (Z)

For the infected asymptomatic, we can use a value of *µ*_1_ = 1*/*11 based on the Singapore study already cited.^9^, alhtough we are aware than other studies such as [25], [9] and [11] have higher estimates, around 20 days.

There is not much information on the variance of the duration in the asymptomatic period and we will use a standard deviation of 7 days.

#### 4.5.3 Hospitalized (H)

This parameter strongly depends on the availability of human resources, equipment and infrastructure. From IMSS data, we estimate it to be 3 days with S.D. of 2.91 days. Thus our estimate of *µ*_3_ = 1*/*3.

#### 4.5.4 Intensive care unit (C)

From IMSS data, we estimate it to be 7 days with S.D. 4 days, for *µ*_4_ = 1*/*7.

#### 4.5.5 Temporary quarantined (R_*T*_)

There is no way to estimate this parameter, since it depends on the time to symptoms onset after the individual has left the *I* stage by self-awareness of testing. We use the memoryless property of exponential distributions. The individual will remain an exponential amount of time in this stage, with parameter equal to the inverse of the average time in the previous stage, which is 1*/*5, but reduce the SD to 4 days.

### 4.6 Estimation of λ and *R*_0_

The baseline *R*_0_ used for the estimations in the Za’atari refugee camp is equal to 3 and is derived from the following equation:

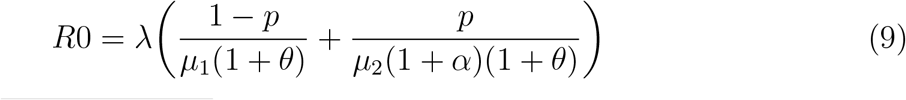

In the equation above, *λ* is an arbitrary parameter which is based on the COVID-19 literature and a specific exercise that we devised to estimate *λ* with publicly available cross-country data. This section explains this exercise and the specific choice of *R*0 = 3 we made.

The contact rate *λ* we attempt to estimate is the *raw contact rate*. That is, it represents the true potential transmission of the disease, and can be measured at the onset of the epidemic. Countries first affected by the pandemic provide a more precise estimate of the pandemic since other countries are expected to be more prepared and are likely to have implemeted measures that may affect *λ*. The *raw contact rate λ* is close to the transmission potential of a disease when the individuals do not react to reduce it, by means of pharmaceutical interventions or by reducing exposure activities. It is important, therefore, to estimate *λ* using existing cross-country data. This will give us information not only on the true potential of the disease but also on how much *λ* can be reduced with preparedness. Once the *λ*s are estimated, we can estimate the respective *R*_0_s.

To estimate *λ*, it is customary to use observations on reported cases, mainly infections, which are recorded mostly when individuals report the onset of symptoms. In our model, that would exclude the *Z* (asymptomatic) individuals and would only consider the *I* (symptomatic) individuals. In most cases, the date reported as the date of a confirmed case is the day a test was made, or even the day the laboratory results were given, and rarely the date of onset of the symptoms. This adds considerable noise to the timing of the cases. Figure 5 shows an hypothetical example of the lag between the time when individuals enter the *I* compartment and the reported time.

As an alternative to using the number of infections, here we use the growth rate in compartment *D* (deaths), which we believe is less prone to error in the timing at which an individual entered that stage. The problem of underreporting is still present but reduced as compared to infections, and the time of death is much more accurate than the time of infection.

In general terms, once all other model’s parameters have been set, we choose an initial *λ* and solve equations 8. With this numerical solution, we obtain the best fit to the observed data in cumulative deaths and then repeat this procedure until the best *λ* value is found within a predefined region. The need to seek the best fit of the solution *D*(*t*) for a given *λ* is due to the fact that the beginning of the outbreak is unknown and thus we need to find a *k* value so that *D*(*t* + *k*) is the best match to the observed data on cumulative deaths.

For this exercise, we use the accumulated number of deaths in 185 countries taken from the data set of Johns Hopkins University [6]. For each country we use the first 15 days of data at the onset of the epidemic starting the day when the accumulated number of deaths was at least 5. Countries with a low *R*^2^ were rejected leaving only 40 countries with reliable estimates. For all countries but Germany, the calculated beginning from our model occurred before the date of first confirmed case.

Table (6) contains the main statistics derived from the cross-country estimation of *λ* and *R*_0_. Estimates of *λ* vary between 0.172 and 0.366 with a mean value of 0.251 with countries that experienced the epidemic at earlier stages showing larger values. From these values of *λ* one can then derive the *R*_0_ as explained above.

**Table 6:**
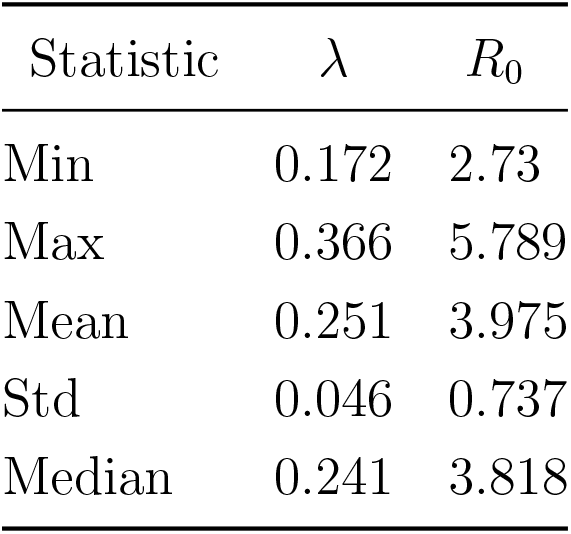
Statistics of the contact rate *λ* and *R*_0_

| Statistic | $\lambda$ | $R_0$ |
| --- | --- | --- |
| Min | 0.172 | 2.73 |
| Max | 0.366 | 5.789 |
| Mean | 0.251 | 3.975 |
| Std | 0.046 | 0.737 |
| Median | 0.241 | 3.818 |

Figure 6 shows the relationship between the age of the pandemic and *R*_0_. For each country, we took as index the day the epidemic reached 5 confirmed cases. Then we took all dates and subtracted the minimum date, so that the indexes are now relative to the country with the first outbreak (excluding China), which is Italy. There is an almost linear relationship between the age of the pandemic and the *R*_0_ in analyzed countries. The fact that *R*_0_ is reduced for countries that experienced the epidemic at later stages indicates the importance and impact of preparing for the epidemic.

In conclusion, the cross-country exercise proposed derived estimates of *λ* that resulted in *R*_0_ varying between 2.73 and 5.789, which are values consistent with the COVID-19 literature. The same exercise also shows that higher values correspond to countries that experienced the epidemic at later stages as compared to Italy. These findings should be taken into account when we consider refugee camps still untouched by the pandemic. We should use a baseline value closer to the lower bound estimated with the cross-country exercise. Hence, our choice of a *R*0 = 3. However, as *R*_0_ is an input parameter, all results can be quickly reproduced with alternative *R*_0_s.

### 4.7 The role of demographics and control policies on the parameters

There are factors affecting outcomes that are characteristics of the demographics of the population. As mentioned before, differential morbidity by age and gender has been amply documented for COVID-19. Also, population size and density and other factors that affect camps specially, like poverty and other health issues may be very important. Control policies like the use of face masks, reducing contacts, contact tracing and testing (*cordon sanitaire*), pharmaceutical treatments, improvement of medical assistance through increasing hospitalization capacity and adding more personnel or even vaccination if it becomes available are all policies that can affect outcomes. In this section, we discuss how all of these changes can be simulated in our model (or are implicit already) without the need of adding more parameters.

1. **Demographics**: The model can accomodate demographic changes as already described in section 4.3 for the estimation of the *adjusted lethality* by age and gender. In essence, demographic changes affect the estimation of *p*, the likelihood that an individual will become a symptomatic infected. As *p* is the first parameter to enter the model, all other estimations take the demographic structure into consideration.
2. **Population density**: this can be included in the contact rate. There is a linear relationship between contact rate and *R*_0_, since this is just the contact rate times the average infectious time. Although we do not exactly know how much the density will affect the contact rate, this effect can be taken into account by multiplying *λ* for a constant (*C*3), which is the number of times we believe the contact rate is increased because of crowdedness.
3. **Household size**: The household size is an important effect to consider, and there are two reasons why we did not include it into the model explicitly. First, household size is implicit in the population density, so, the larger the household size, the larger the density per square km, and thus we can increase the contact rate, as pointed out before. Second, in the particular case of COVID-19, which is highly contagious, if we are willing to assume that all members of the household will be infected when a single member of the household becomes infected, then it is possiblle to show that *R*_0_ increases by a factor equal to the average household size, which will take the epidemic size to a much higher level. Since, as it will be discussed later, we can expect a very large fraction of the population to become infected without modeling households, adding this level of complexity is unnecessary.
4. **Health issues** There are several studies on how the lethality of SARS-CoV-2 is increased due to health issues like obesity, diabetes, respiratory diseases or habits like smoking [18, 22, 1]. Most odds ratios in those studies are large with a large SD. Including this differential lethality in our model for RC is impossible because there is no information available on the incidence of these conditions among the refugees. We can only think that the likelihood of becoming symptomatic (parameter *p*), the chances of moving to ICU while in hospitalization (parameter *q*), and the chances of dying while in ICU (parameter *w*) in our model all increase if health issues are present. However, the amount is left to speculation.
5. **Poverty** Poverty includes a wide range of factors that affect the outcome of the epidemic in a given community. Poverty is related to reduced education (which affects the ability of camp administrators to implement control measures), diminished nutrition and reduced medical attention. There are no specific studies that analyze how poverty affects the outcome of an epidemic but surely a reduced level of education tends to reduce the perception of the importance of the disease and thus may not reduce the contact rate *λ*, whereas lack of nutrition and medical attention affect *p, q* and *w* in our model. Again, the amount to increase is left to speculation. An analysis of the relationship between the number of doctors in a city as a surrogate of wealth, and the number of cases is shown in [19].
6. **Social distancing and lock-down** Social distancing is reflected in the model as a reduction in the contact rate *λ*. In Figure 6 we saw how the *R*_0_ is reduced in time across countries due to learning, which allowed some countries to prepare for the arrival of the pandemic. This implies that it is possible to prepare by reducing the contact rate through reinforcing and encouraging social distancing. However, in refugee camps measures such as social distancing are evidently more difficult to apply. Also, the possibility that the disease will not enter a refugee camp is very low as camps’ supplies strongly depend on the interaction with people outside the camp.
7. **Face masks** The effect of face masks is similar to that of an *altruistic* vaccine. It is not very effective in reducing infection of the mask-user, but reduces the ability to transmit the infection. Therefore, regarding the epidemic size, they act as a vaccine. The use of surgical masks has been reported to be effective in the transmission of the disease [7, 5], and [12] even suggests that frequent mask use in public venues, frequent hand washing, and disinfecting the living quarters were significant protective factors of infections (Odds Ratio 0.36 to 0.58). Nevertheless, the large number of infections reported among nurses and doctors that certainly worked wearing a surgical mask, imply some limit to this protective effect in crowded environments with a lot of infected people. If wearing a mask reduces the emission of coronavirus droplets from an infected person to a fraction *φ*, then lambda is reduced to *λφ*. If we assume that masks protect equally susceptible persons by reducing the virus droplets by the same amount, then *λ* is reduced to *λφ*^2^. There is no information on the value of *φ*, and we only can speculate on some positive value. Nevertheless, we must be cautious since by considering that *λ* reduces to *λφ*^2^ we are implicitly assuming that a reduction in virus droplets by a half will reduce the likelihood of being infected by a half, which is not necessarily true.
8. **Contact tracing, testing and quarantine** As mentioned earlier, the *cordon sanitaire* consists on locating people that had contact with an infected individual and quarantine them. Parameter *θ* in our model is an attempt to account for this effect, in the sense that *θ* is how good we are at detecting and isolating infected contacts. However, *θ* is a parameter we need to input as a guess estimate.
9. **Vaccination** A vaccine that has efficacy *β* reduces the contact rate to *λβ*. There is no current candidate vaccine and is seriously challenged that it will be developed within a year. Even if a candidate vaccine is available now, the testing time through phase IV takes over a year, even if all the permits are speeded up. Therefore, modelling the impact of a vaccine is currently premature.
10. **Pharmaceutical interventions** Pharmaceutical interventions or any other treatment is included in parameters *q* and *w* in the model. Any increase in survival of individuals due to medical treatment should be included as a decrease in *q* or *w*.
11. **Herd immunity** The herd immunity is just the *thinning* term *S/N* in the first two rows of Table 2. The larger the infected group, the smaller the probability that the next contact of an infected person will be with a susceptible person. The highest peak of herd immunity is reached when the product (*I* +*Z*)*∗S/N* is maximum, which is achieved when the number of active infected is equal to the number of susceptible. Although sometimes herd immunity has been referred to as a policy or resource, this is not the case. Exposing the population by not implementing social distancing or lock-down will only accelerate the time to final outcome and will increase the demand of medical services at any time.

There has been some claims that there is no guarantee that immunity is achieved. Still, the number of cases reported at the global level of a second infection are scarce and dubious. If immunity is a fact, then, surveillance for immunity is useful in order to avoid a full collapse of economic activities in the camp. Random testing, and in particular testing those who recovered, is important to measure the size of compartment *R* at all times. There is the possibility of deducting this by analyzing the ratio Recovered/Deaths. At the worldwide level, on May 19, JHU reported 319, 213 deaths and 1, 805, 093 recovered, that is, for every death there are at least 5.6 recovered. This is a very conservative estimate, because the number of undetected recoveries is larger than the number of undetected deaths, since most of the recoveries detected were more among symptomatic than among asymptomatic, and the first group has a larger mortality than the second.

### 4.8 Sensitivity analysis

A sensitivity analysis consists in analyzing the effect of changes in the parameters on the output of the epidemic. We concentrate on the effect of the parameters on *R*_0_. With *L*_*Z*_ = 1*/µ*_1_, *L*_*I*_ = 1*/µ*_2_ where *L*_*I*_ and *L*_*Z*_ are the respective average times in compartments *I* and *Z*, the equation for *R*_0_ can be expressed as follows:

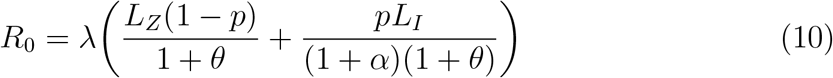

We can see in (10) that *R*_0_ depends on *λ, p, µ*_1_, *µ*_2_, *θ* and *α*. As mentioned before, the value of *p* is determined by the nature of the disease and the demographic structure of the population and, in this sense, *p* can be considered as a fixed and exogenous parameter. Changes in *λ* are possible by reducing the contact rate with active policies such as social distancing or the use of face masks [7, 5, 12]. Changes in *µ*_1_ and *µ*_2_ are possible by testing and isolation policies for the infected, which reduce the average time in compartments *Z* and *I*, and increase the exit rates from these compartments.

How sensitive is *R*_0_ to changes in *λ, θ* and *α*? Figure 4 shows changes in *R*_0_ when *λ* = 0.3, *L*_*I*_ = 5, *L*_*Z*_ = 11, *p* = 0.0172, *θ* = 0.25 and *α* = 0.25. The left-hand panel shows absolute changes whereas the right-hand panel shows percentage changes (elasticities). Results show that *λ* is the most elastic and important parameter (a 1% increase in *λ* resuts in a 1% increase in *R*_0_), which implies that we need to focus on reducing the contact rate. *θ* and *α* are much less elastic, implying that it is much more difficult to contain the spread of the disease with testing, contact tracing or self-isolation due to symptom awareness. This is mainly due to the fact that there is a smaller amount of symptomatic individuals that could reduce their infectious time and have a significant effect on the epidemic.

**Figure 3:**
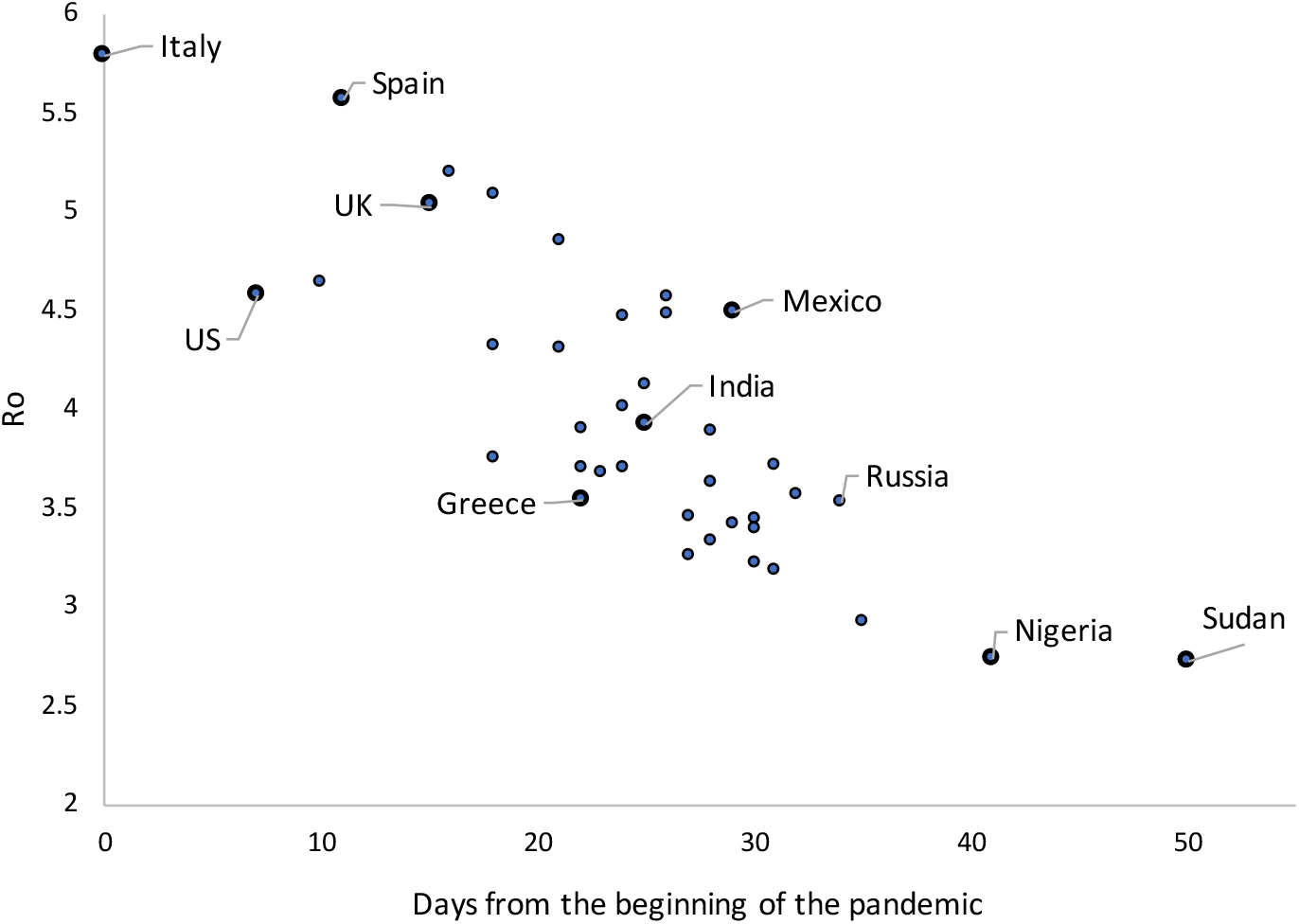
Age of the pandemic vs. *R*_0_. For each country, the baseline was taken as the date when the accumulated number of deaths was at least 5. Italy is the baseline on 02/24/2020.

**Figure 4:**
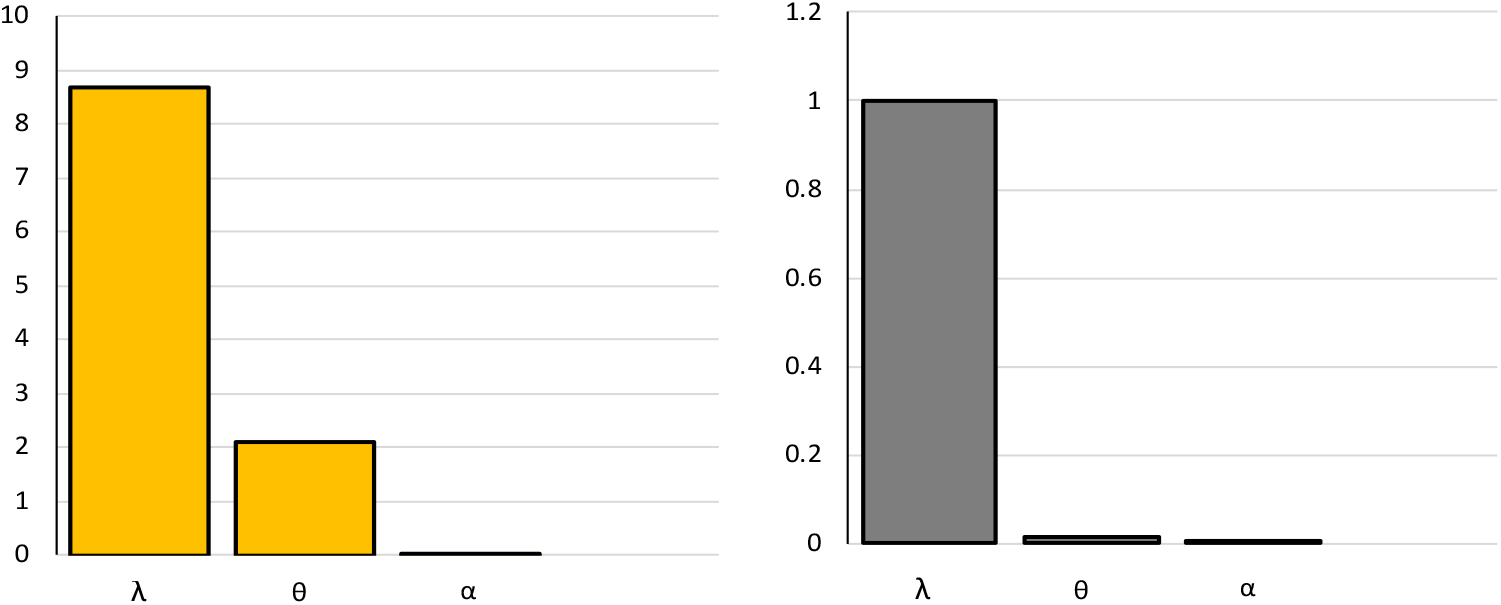
Sensitivity analysis of *R*_0_ to *λ, θ* and *α*. Absolute values (left panel) and elasticities (right-panel).

### 4.9 Summary of parameters for the Za’atari camp estimations

Table 7 summarizes the values of the parameters discussed in this report and used for the estimation of the SIZ model using 2015 data for the Za’atari refugee camp.

**Table 7:** Parameters summary

| Parameter description | Symbol | Value |
| --- | --- | --- |
| Basic reproductive number | $R_0$ | 3 |
| Proportion of infected that will be hospitalized | p | 0.0026 |
| Proportion of hospitalized individuals that requires intensive care | q | 0.25 |
| Proportion of individuals in intensive care that will die | w | 0.67 |
| Proportion of infected removed due to testing and contact tracing | $\theta$ | varies |
| Proportion of voluntary quarantine due to self-awareness | $\alpha$ | 0 |
| Time in Z | $1/\mu_1$ | 11 |
| Time in I | $1/\mu_2$ | 5 |
| Time in RT | $1/\mu_2$ | 5 |
| Time in H | $1/\mu_3$ | 3 |
| Time in C | $1/\mu_4$ | 7 |
| Reduction in contact rate by social distance | C1 | varies |
| Reduction in contact rate by use of mask | C2 | 0 |
| Increase severity for camp conditions | C3 | 3 |

## 5 The Stochastic Solution of the Model

In a stochastic model, the current state of the process is a vector **X**(*t*_0_) that will change to some state **X**(*t*_0_ + *t*) in some time *t*. Whereas in deterministic models the word *simulation* is used to refer to the *numerical solution* of a system of differential equations, in stochastic models it has a different interpretation: the transition from the current state **X**(*t*_0_) to the state **X**(*t*_0_ + *t*) is random, as well as the time for the transition, *t*. Given some current state of the process, the change in the state that will occur next as well as when the change will take place are calculated using *transition rates* expressed in terms of probabilities.

The main difference between deterministic and stochastic models reside in the algorithms for the transitions between compartments. Suppose that, after leaving a compartment, a person can move to two different compartments at rates *α*_1_ and *α*_2_ respectively. In a deterministic model, a fraction *α*_1_*/*(*α*_1_ + *α*_2_) moves to one compartment and another fraction *α*_2_*/*(*α*_1_ + *α*_2_) moves to the other. In a stochastic model, the individual moves to each compartment according to probabilities. If we take a look at the model in Figure 1, we can see for instance that an individual in stage *H* can move to stage *C* or *R* at rates *µ*_3_*q* and *µ*_3_(1 *– q*) respectively. Therefore, the probability that this person will move to *C* is *µ*_3_*q/*(*µ*_3_*q* + *µ*_3_(1 *– q*)) = *q*, which is the relative share of those rates. To decide if the individual moves to *R* or *C* we generate a random number *u* from a uniform distribution in (0, 1) and if *u < q* the individual moves to stage *C*, otherwise this individual moves to *R*. In the deterministic model, a fraction *q* of the individual moves to stage *C* and a fraction 1 *– q* to stage *R*.

Given the current state of the process as a vector containing the number of individuals in each class of:

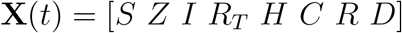

we first construct a list of all possible events that can take place at the next step, along with their rates. In our particular model, this list is given in Table 8. Observe that the list is obtained from the rates of the differential equations model. To decide which of the 12 possible events will take place, we take a single observation from a multinomial random variable with parameters

**Table 8:** List of all possible events and their rates

| From: | To: | Event number | Transition Rates |
| --- | --- | --- | --- |
| $Z$ | $Z + 1$ | 1 | $(1 - p)\lambda S(I + Z)/N$ |
| | $Z - 1$ | 2 | $\mu_1 Z(1 + \theta)$ |
| $I$ | $I + 1$ | 3 | $p\lambda S(I + Z)/N$ |
| | $I - 1$ | 4 | $\mu_2(1 + \alpha)(1 + \theta)I$ |
| $R_T$ | $R_T + 1$ | 5 | $\mu_2(\alpha + \theta(2 + \alpha))I$ |
| | $R_T - 1$ | 6 | $\mu_2 R_T$ |
| $H$ | $H + 1$ | 7 | $\mu_2 R_T + \mu_2(1 - \theta)I$ |
| | $H - 1$ | 8 | $\mu_3 H$ |
| $C$ | $C + 1$ | 9 | $\mu_3 q H$ |
| | $C - 1$ | 10 | $\mu_4 C$ |
| $R$ | $R + 1$ | 11 | $(1 - q)\mu_3 H + (1 - w)\mu_4 C + \mu_1(1 + \theta)Z$ |
| $D$ | $D + 1$ | 12 | $\mu_4 w C$ |

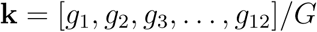

where *g*_*i*_ = rate of event and *G* = ∑_*i*_ *g*_*i*_. Once we decide which event takes place, we update the number of individuals in each compartment and recalculate the rates, then update parameter **k** in the multinomial distribution and sample it again until ∑_*i*_ *g*_*i*_ = 0 or up to a specific time *t*. Table 8 provides all possible transitions with the corresponding rates.

### 5.1 Time to event simulation

In the previous section we explained how to simulate a series of transitions in a continuous-time discrete-state stochastic process, that produces a vector **X**(*t*) at each iteration. Nevertheless, we need to simulate the time between transitions, that is, the index *t*.

In a stochastic model, when an individual is in some compartment and this individual leaves the compartment at a rate *µ*, it means that the individual will remain in that compartment according to a random variable *T* that follows an exponential distribution with parameter *µ*. That is:

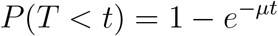

If there are *n* individuals in that compartment, the time for the next individual to leave the compartment is clearly the minimum of *n* exponential random variables, which is also exponential with parameter *nµ*, that is, the time for the next individual to leave the compartment is:

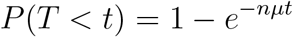

Therefore, we can simulate the time to the next individual to leave this compartment using:

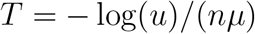

where *u* is a random number from an uniform distribution in (0, 1). Using this, the t e to the next transition is an exponential random variable with parameter *R* = ∑_*i*_ *R*_*i*_. Therefore, the time to the next transition can be simulated as:

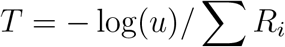

It is important to notice that the inter-event times are independent of the event that will take place. That is, the time to the next transition and the type of the next transition are independent.

### 5.2 Accelerating the simulations

*Time to event* simulations are generally inefficient computationally since only one individual moves at the time. The simulation stops when no more individuals can leave a compartment which in our case is when all individuals are distributed among *S, R* or *D*. The number of transitions depends largely on the population size, and for a population of size *N* it could be up to 5*N*, since in our model approximately *N* individuals will move to up to 5 stages.

There is a way to speed up the simulations, based on the relationship between exponential and Poisson distributions. This can be explained by looking at Figure 5. In this figure, there are *n* = 8 events that take place at the same rate. In *time to event* simulations the system will change state when the first event occurs, which is event *E*_5_ in Figure 5. In our approximation, we select a window of size *W*, and calculate the probability that an event takes place in that window, call this *p*. Then, the number of events that takes place in that window is a Binomial random variable with parameters (*n, p*). Since the number of individuals in a compartment is, in general, large and *W* can be very small, we can then approximate the binomial distribution with a Poisson distribution with parameter *np*. Clearly, the limit when *W →* 0 is the same as performing *time to event* simulation. In short, instead of simulating one single event at the time, we generate all the events that will occur in a window of size *W*. The smaller the size of the window, the better the approximation, because the events inside *W* are not independent.

In our simulations, we take *W* = 1*/*24, that is, we change the system at unit steps of an hour. This will require about 365 *×* 24 = 8, 760 transitions to simulate an outbreak with a duration of one year, instead of the approximate half a million simulations for a refugee camp of 100, 000 people. Another advantage is that this Poisson approximation is independent of the population size.

### 5.3 Matching the mean and variance in stochastic simulations

Next, we calibrate simulations to reproduce real data. The idea is to match the reported (observed) mean and variance of different stages, for instance, for the *Z* and *I* stages. Assuming that the time in a compartment is exponential yields a mean equal to the standard deviation, which is undesirable. For instance, if we assume that the time in stage *Z* lasts on average 11 days, then an ordinary stochastic approach to the model in Figure 1 will cause an individual to stay in that stage for an average 11 days with a standard deviation of 11 days, which is different from the reported (observed) 7 days that can be found in the literature and are described in the deterministic report.

The method consits of dividing each stage in several compartments with an exponential duration each, so that the passage through all of them has the desired mean and variance. The number of compartments and the individual rates depend on how precise is the approximation we want to those two first central moments. This method of approximation is called *Phase distribution*. [16, 17, 3, 14, 4]. In theory, it is possible to match as many moments as we want, although in here we only try to match the first two central moments, and we sacrifice precision in order to keep the number of total compartments relatively small. Observe that our method is different from the one reviewed in [4] in which a stage is subdivided in compartments with identical exit rates. Phase distribution methods are not restricted to identical exit rates.

Figure 6 shows the number of compartments required for each stage, Table 9 shows the required rates for each compartment and Table 10 shows the match achieved between the mean and variance reported in the literature and the one achieved with our approximation.

**Table 9:** Rates in Figure 6

| Stage | Exit rates |
| --- | --- |
| $Z$ | $r_{11} = (1 + \theta)1.833^{-1}$ |
| $C$ | $r_{21} = 2.14^{-1}, r_{22} = 1.359^{-1}$ |
| $H$ | $r_{31} = 1.899^{-1}, r_{32} = 0.983^{-1}$ |
| $I$ | $r_{41} = (1 + \theta)(1 + \alpha) 2.5^{-1}$ |
| $R_T$ | $r_{51} = 3^{-1}, r_{52} = 2^{-1}$ |

**Table 10:** Moments and their match in the stochastic model (days)

| Parameter | Observed |  | Model |  |
| --- | --- | --- | --- | --- |
|  | Mean | SD | Mean | SD |
| Time in $Z$ | 11 | 7 | 11 | 5.8 |
| Time in $I$ | 5 | 4.3 | 5 | 3.73 |
| Time in $R_T$ | 5 | 4 | 5 | 3.6 |
| Time in $H$ | 3 | 2.91 | 2.88 | 2.14 |
| Time in $C$ | 7 | 4 | 7 | 3.59 |

## Data Availability

No data available

https://bit.ly/3jdtOXX

## 6 Acknowledgments

The report is part of the DFID-UNHCR-World Bank program “Building the Evidence on Protracted Forced Displacement: A Multi-Stakeholder Partnership”. The program is funded by UK aid from the United Kingdom’s Department for International Development (DFID), it is managed by the World Bank Group (WBG) and was established in partnership with the United Nations High Commissioner for Refugees (UNHCR). The scope of the program is to expand the global knowledge on forced displacement by funding quality research and disseminating results for the use of practitioners and policy makers. This work does not necessarily reflect the views of DFID, the WBG or UNHCR. We are grateful to Ann Burton (UNHCR) and Paul Spiegel (John Hopkins University) for comments and clarifications.

## A Results - Deterministic model

**Table 11:** Summary of simulations

| Contact trace<br>policy ($\theta$)* | Contact rate reduction<br>policy ($C_1$) | Table: |
| --- | --- | --- |
| No contact tracing | No reduction in contact rate | Table 12 |
|  | 33 % reduction in contact rate | Table 13 |
| Contact tracing: 33 % | No reduction in contact rate | Table 14 |
|  | 33 % reduction in contact rate | Table 15 |

\*The fraction of the infected contacts that are traced, tested and quarantined.

**Table 12:**
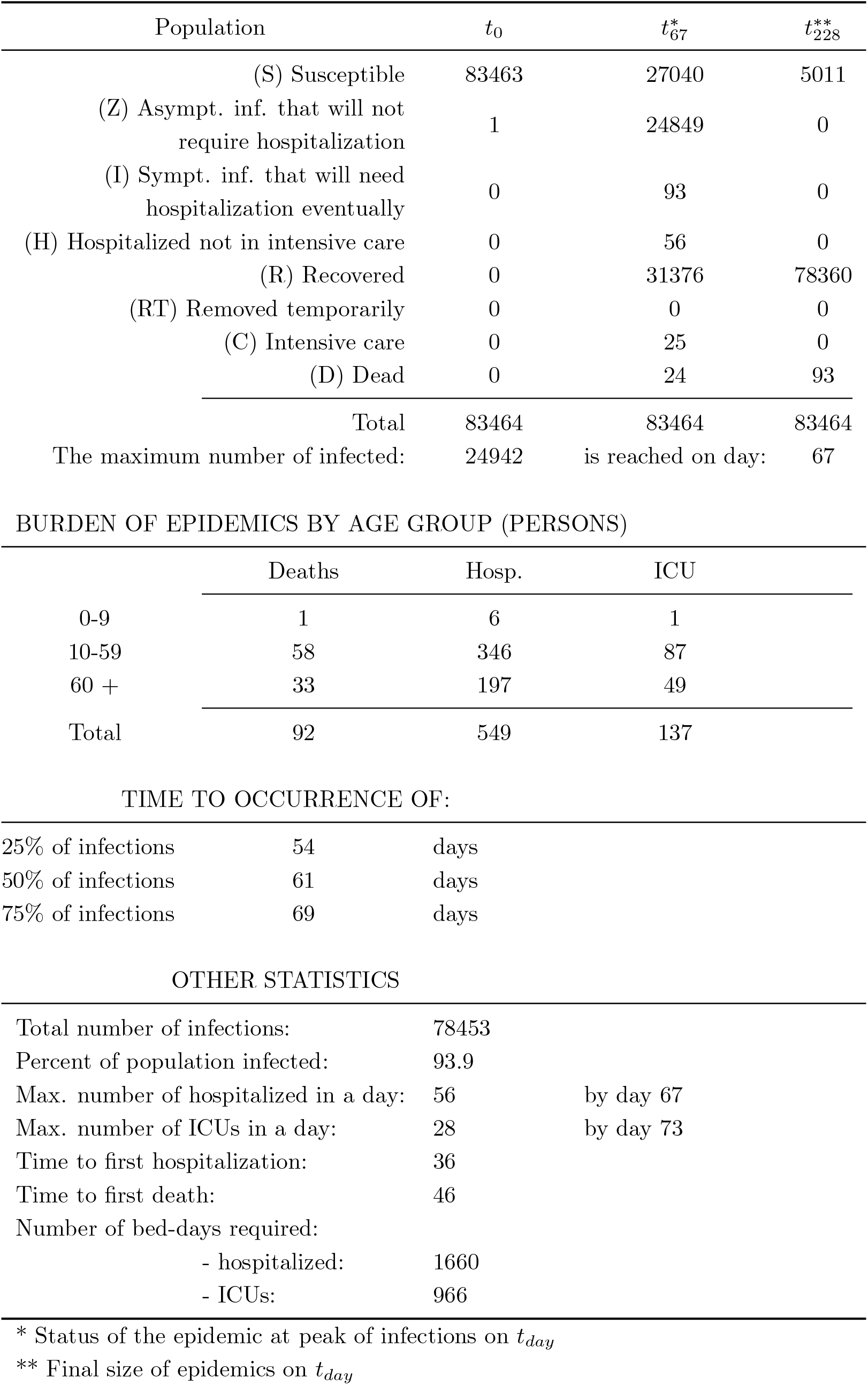
SIZ Model: No reduction in contact rate and no contact tracing.

| Population | $t_0$ | $t_{67}^*$ | $t_{228}^{**}$ |
| --- | --- | --- | --- |
| (S) Susceptible | 83463 | 27040 | 5011 |
| (Z) Asympt. inf. that will not<br>require hospitalization | 1 | 24849 | 0 |
| (I) Sympt. inf. that will need<br>hospitalization eventually | 0 | 93 | 0 |
| (H) Hospitalized not in intensive care | 0 | 56 | 0 |
| (R) Recovered | 0 | 31376 | 78360 |
| (RT) Removed temporarily | 0 | 0 | 0 |
| (C) Intensive care | 0 | 25 | 0 |
| (D) Dead | 0 | 24 | 93 |
| Total | 83464 | 83464 | 83464 |
| The maximum number of infected: | 24942 | is reached on day: | 67 |

BURDEN OF EPIDEMICS BY AGE GROUP (PERSONS)
|  | Deaths | Hosp. | ICU |
| --- | --- | --- | --- |
| 0-9 | 1 | 6 | 1 |
| 10-59 | 58 | 346 | 87 |
| 60 + | 33 | 197 | 49 |
| Total | 92 | 549 | 137 |

TIME TO OCCURRENCE OF:
|  |  |  |
| --- | --- | --- |
| 25% of infections | 54 | days |
| 50% of infections | 61 | days |
| 75% of infections | 69 | days |

OTHER STATISTICS
|  |  |  |
| --- | --- | --- |
| Total number of infections: | 78453 |  |
| Percent of population infected: | 93.9 |  |
| Max. number of hospitalized in a day: | 56 | by day 67 |
| Max. number of ICUs in a day: | 28 | by day 73 |
| Time to first hospitalization: | 36 |  |
| Time to first death: | 46 |  |
| Number of bed-days required: |  |  |
| - hospitalized: | 1660 |  |
| - ICUs: | 966 |  |

\* Status of the epidemic at peak of infections on $t_{day}$ \*\* Final size of epidemics on $t_{day}$

**Table 13:**
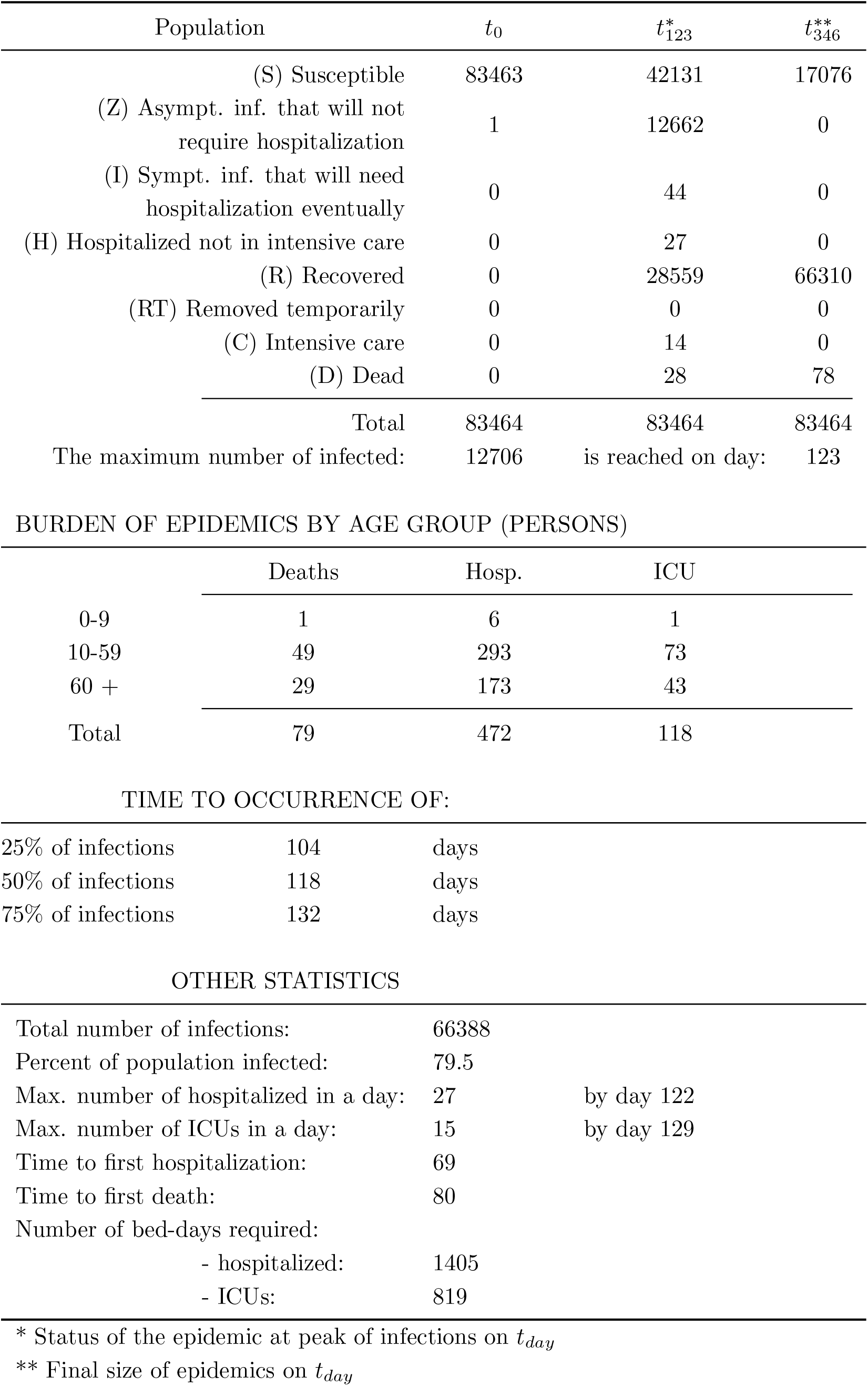
SIZ Model: 1/3 reduction in contact rate and no contact tracing.

| Population | $t_0$ | $t_{123}^*$ | $t_{346}^{**}$ |
| --- | --- | --- | --- |
| (S) Susceptible | 83463 | 42131 | 17076 |
| (Z) Asympt. inf. that will not<br>require hospitalization | 1 | 12662 | 0 |
| (I) Sympt. inf. that will need<br>hospitalization eventually | 0 | 44 | 0 |
| (H) Hospitalized not in intensive care | 0 | 27 | 0 |
| (R) Recovered | 0 | 28559 | 66310 |
| (RT) Removed temporarily | 0 | 0 | 0 |
| (C) Intensive care | 0 | 14 | 0 |
| (D) Dead | 0 | 28 | 78 |
| Total | 83464 | 83464 | 83464 |
| The maximum number of infected: | 12706 | is reached on day: | 123 |

BURDEN OF EPIDEMICS BY AGE GROUP (PERSONS)
|  | Deaths | Hosp. | ICU |
| --- | --- | --- | --- |
| 0-9 | 1 | 6 | 1 |
| 10-59 | 49 | 293 | 73 |
| 60 + | 29 | 173 | 43 |
| Total | 79 | 472 | 118 |

TIME TO OCCURRENCE OF:
|  |  |  |
| --- | --- | --- |
| 25% of infections | 104 | days |
| 50% of infections | 118 | days |
| 75% of infections | 132 | days |

OTHER STATISTICS
|  |  |  |
| --- | --- | --- |
| Total number of infections: | 66388 |  |
| Percent of population infected: | 79.5 |  |
| Max. number of hospitalized in a day: | 27 | by day 122 |
| Max. number of ICUs in a day: | 15 | by day 129 |
| Time to first hospitalization: | 69 |  |
| Time to first death: | 80 |  |
| Number of bed-days required: |  |  |
| - hospitalized: | 1405 |  |
| - ICUs: | 819 |  |

\* Status of the epidemic at peak of infections on $t_{day}$ \*\* Final size of epidemics on $t_{day}$

**Table 14:**
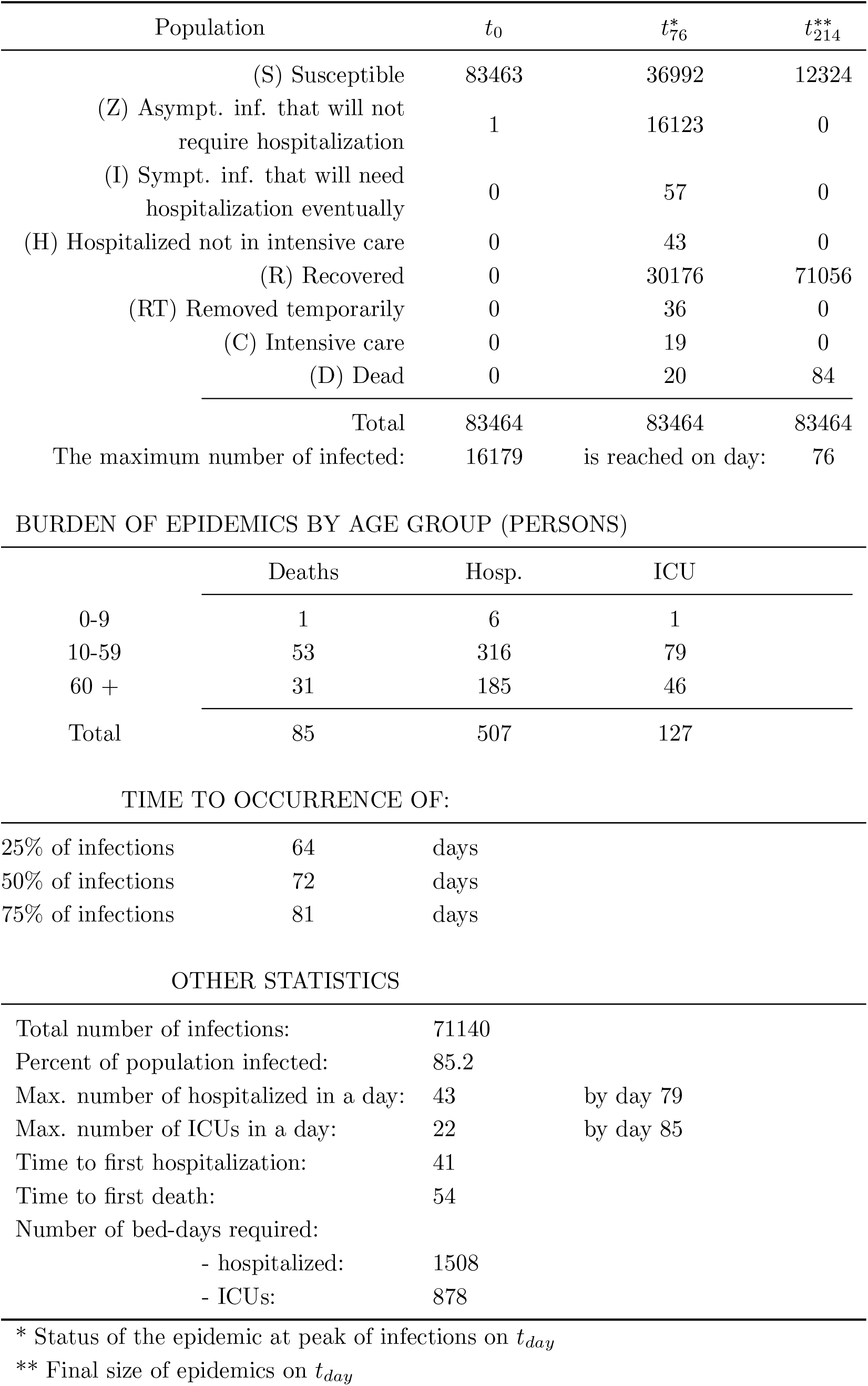
SIZ Model: no reduction in contact rate and contact tracing *θ* = 1*/*3.

| Population | $t_0$ | $t_{76}^*$ | $t_{214}^{**}$ |
| --- | --- | --- | --- |
| (S) Susceptible | 83463 | 36992 | 12324 |
| (Z) Asympt. inf. that will not<br>require hospitalization | 1 | 16123 | 0 |
| (I) Sympt. inf. that will need<br>hospitalization eventually | 0 | 57 | 0 |
| (H) Hospitalized not in intensive care | 0 | 43 | 0 |
| (R) Recovered | 0 | 30176 | 71056 |
| (RT) Removed temporarily | 0 | 36 | 0 |
| (C) Intensive care | 0 | 19 | 0 |
| (D) Dead | 0 | 20 | 84 |
| Total | 83464 | 83464 | 83464 |
| The maximum number of infected: | 16179 | is reached on day: | 76 |

BURDEN OF EPIDEMICS BY AGE GROUP (PERSONS)
|  | Deaths | Hosp. | ICU |
| --- | --- | --- | --- |
| 0-9 | 1 | 6 | 1 |
| 10-59 | 53 | 316 | 79 |
| 60 + | 31 | 185 | 46 |
| Total | 85 | 507 | 127 |

TIME TO OCCURRENCE OF:
|  |  |  |
| --- | --- | --- |
| 25% of infections | 64 | days |
| 50% of infections | 72 | days |
| 75% of infections | 81 | days |

OTHER STATISTICS
|  |  |  |
| --- | --- | --- |
| Total number of infections: | 71140 |  |
| Percent of population infected: | 85.2 |  |
| Max. number of hospitalized in a day: | 43 | by day 79 |
| Max. number of ICUs in a day: | 22 | by day 85 |
| Time to first hospitalization: | 41 |  |
| Time to first death: | 54 |  |
| Number of bed-days required: |  |  |
| - hospitalized: | 1508 |  |
| - ICUs: | 878 |  |

\* Status of the epidemic at peak of infections on $t_{day}$ \*\* Final size of epidemics on $t_{day}$

**Table 15:**
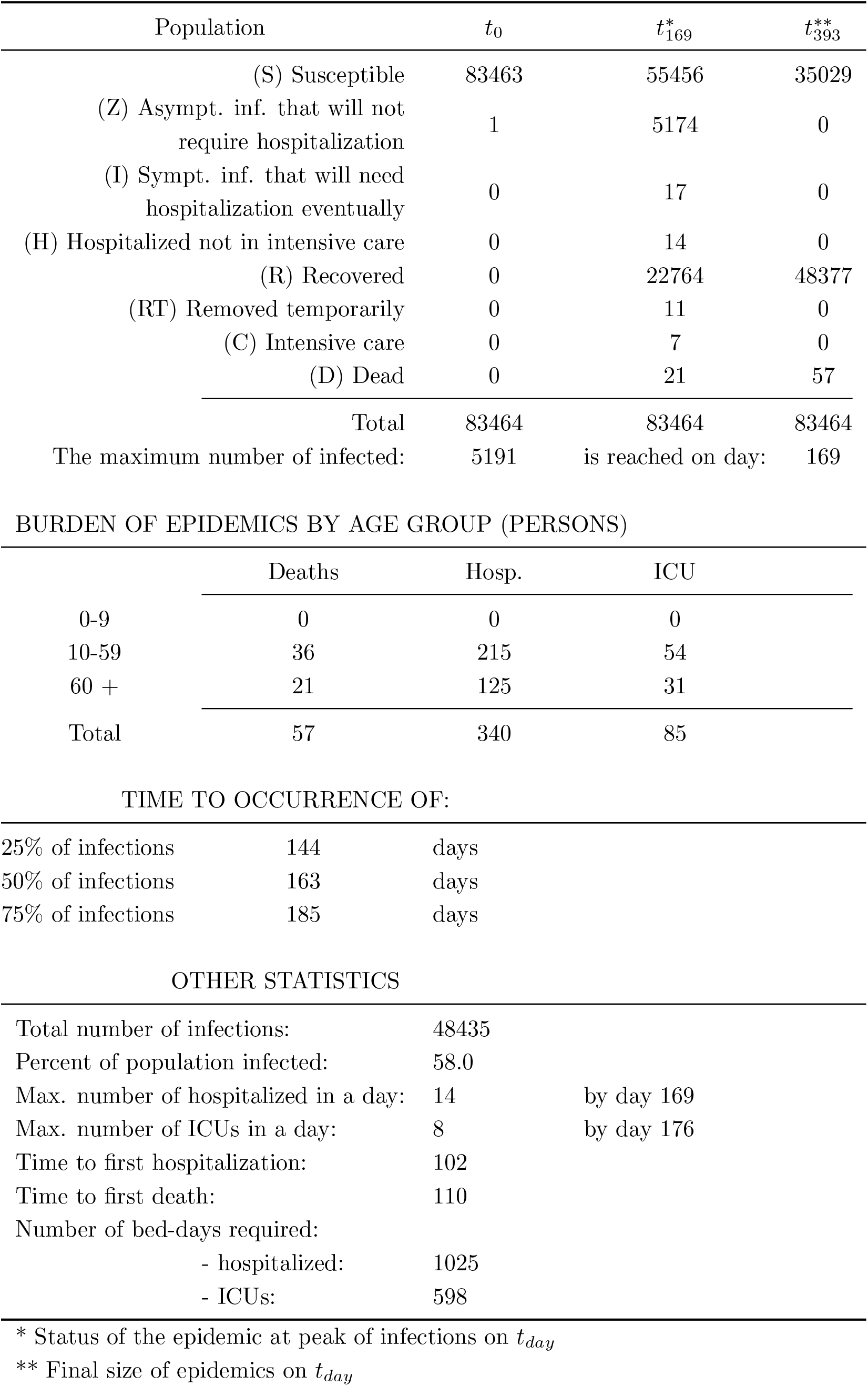
SIZ Model: 1*/*3 reduction in contact rate and contact tracing *θ* = 1*/*3.

| Population | $t_0$ | $t_{169}^*$ | $t_{393}^{**}$ |
| --- | --- | --- | --- |
| (S) Susceptible | 83463 | 55456 | 35029 |
| (Z) Asympt. inf. that will not<br>require hospitalization | 1 | 5174 | 0 |
| (I) Sympt. inf. that will need<br>hospitalization eventually | 0 | 17 | 0 |
| (H) Hospitalized not in intensive care | 0 | 14 | 0 |
| (R) Recovered | 0 | 22764 | 48377 |
| (RT) Removed temporarily | 0 | 11 | 0 |
| (C) Intensive care | 0 | 7 | 0 |
| (D) Dead | 0 | 21 | 57 |
| Total | 83464 | 83464 | 83464 |
| The maximum number of infected: | 5191 | is reached on day: | 169 |

BURDEN OF EPIDEMICS BY AGE GROUP (PERSONS)
|  | Deaths | Hosp. | ICU |
| --- | --- | --- | --- |
| 0-9 | 0 | 0 | 0 |
| 10-59 | 36 | 215 | 54 |
| 60 + | 21 | 125 | 31 |
| Total | 57 | 340 | 85 |

TIME TO OCCURRENCE OF:
|  |  |  |
| --- | --- | --- |
| 25% of infections | 144 | days |
| 50% of infections | 163 | days |
| 75% of infections | 185 | days |

OTHER STATISTICS
|  |  |  |
| --- | --- | --- |
| Total number of infections: | 48435 |  |
| Percent of population infected: | 58.0 |  |
| Max. number of hospitalized in a day: | 14 | by day 169 |
| Max. number of ICUs in a day: | 8 | by day 176 |
| Time to first hospitalization: | 102 |  |
| Time to first death: | 110 |  |
| Number of bed-days required: |  |  |
| - hospitalized: | 1025 |  |
| - ICUs: | 598 |  |

\* Status of the epidemic at peak of infections on $t_{day}$ \*\* Final size of epidemics on $t_{day}$

## B Results - Stochastic Model

**Table 16:** Summary of simulations

| Contact trace<br>policy ($\theta$)* | Contact rate reduction<br>policy ($C_1$) | Table: |
| --- | --- | --- |
| No contact tracing | No reduction in contact rate | Table 17 |
|  | 33 % reduction in contact rate | Table 18 |
| Contact tracing: 33 % | No reduction in contact rate | Table 19 |
|  | 33 % reduction in contact rate | Table 20 |

\*The fraction of the infected contacts that is traced, tested and quarantined.

**Table 17:**
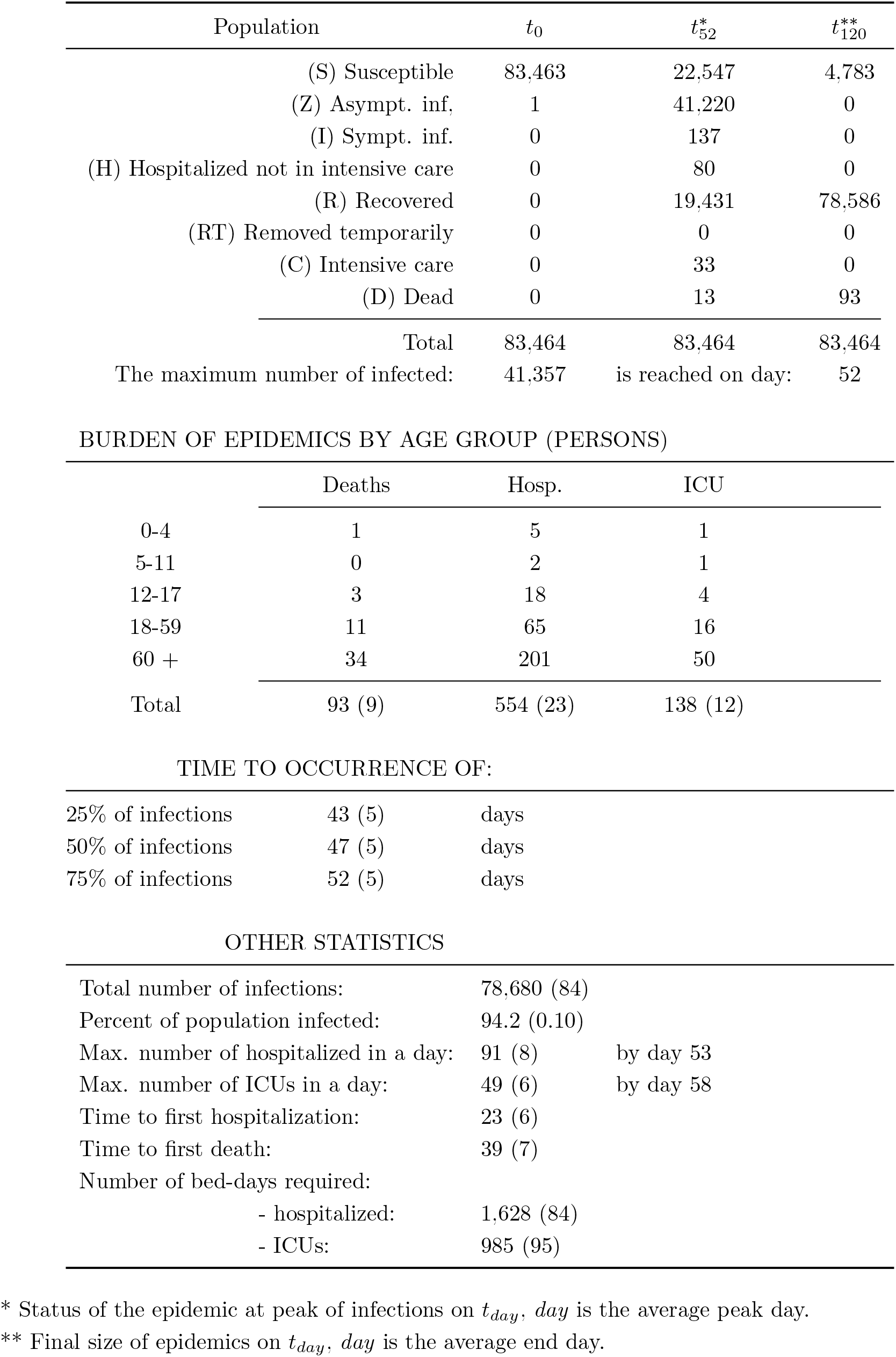
SIZ Model: *R*_0_ = 3, *θ* = 0, *C*_1_ = 0.0, *C*_2_ = 0.0, *C*_3_ = 3 (Numbers in parenthesis are SD)

| Population | $t_0$ | $t_{52}^*$ | $t_{120}^{**}$ |
| --- | --- | --- | --- |
| (S) Susceptible | 83,463 | 22,547 | 4,783 |
| (Z) Asympt. inf, | 1 | 41,220 | 0 |
| (I) Sympt. inf. | 0 | 137 | 0 |
| (H) Hospitalized not in intensive care | 0 | 80 | 0 |
| (R) Recovered | 0 | 19,431 | 78,586 |
| (RT) Removed temporarily | 0 | 0 | 0 |
| (C) Intensive care | 0 | 33 | 0 |
| (D) Dead | 0 | 13 | 93 |
| Total | 83,464 | 83,464 | 83,464 |
| The maximum number of infected: | 41,357 | is reached on day: | 52 |

BURDEN OF EPIDEMICS BY AGE GROUP (PERSONS)
|  | Deaths | Hosp. | ICU |
| --- | --- | --- | --- |
| 0-4 | 1 | 5 | 1 |
| 5-11 | 0 | 2 | 1 |
| 12-17 | 3 | 18 | 4 |
| 18-59 | 11 | 65 | 16 |
| 60 + | 34 | 201 | 50 |
| Total | 93 (9) | 554 (23) | 138 (12) |

TIME TO OCCURRENCE OF:
|  |  |  |
| --- | --- | --- |
| 25% of infections | 43 (5) | days |
| 50% of infections | 47 (5) | days |
| 75% of infections | 52 (5) | days |

OTHER STATISTICS
|  |  |  |
| --- | --- | --- |
| Total number of infections: | 78,680 (84) |  |
| Percent of population infected: | 94.2 (0.10) |  |
| Max. number of hospitalized in a day: | 91 (8) | by day 53 |
| Max. number of ICUs in a day: | 49 (6) | by day 58 |
| Time to first hospitalization: | 23 (6) |  |
| Time to first death: | 39 (7) |  |
| Number of bed-days required: |  |  |
| - hospitalized: | 1,628 (84) |  |
| - ICUs: | 985 (95) |  |

\* Status of the epidemic at peak of infections on $t_{day}$ , $day$ is the average peak day.\*\* Final size of epidemics on $t_{day}$ , $day$ is the average end day.

**Table 18:**
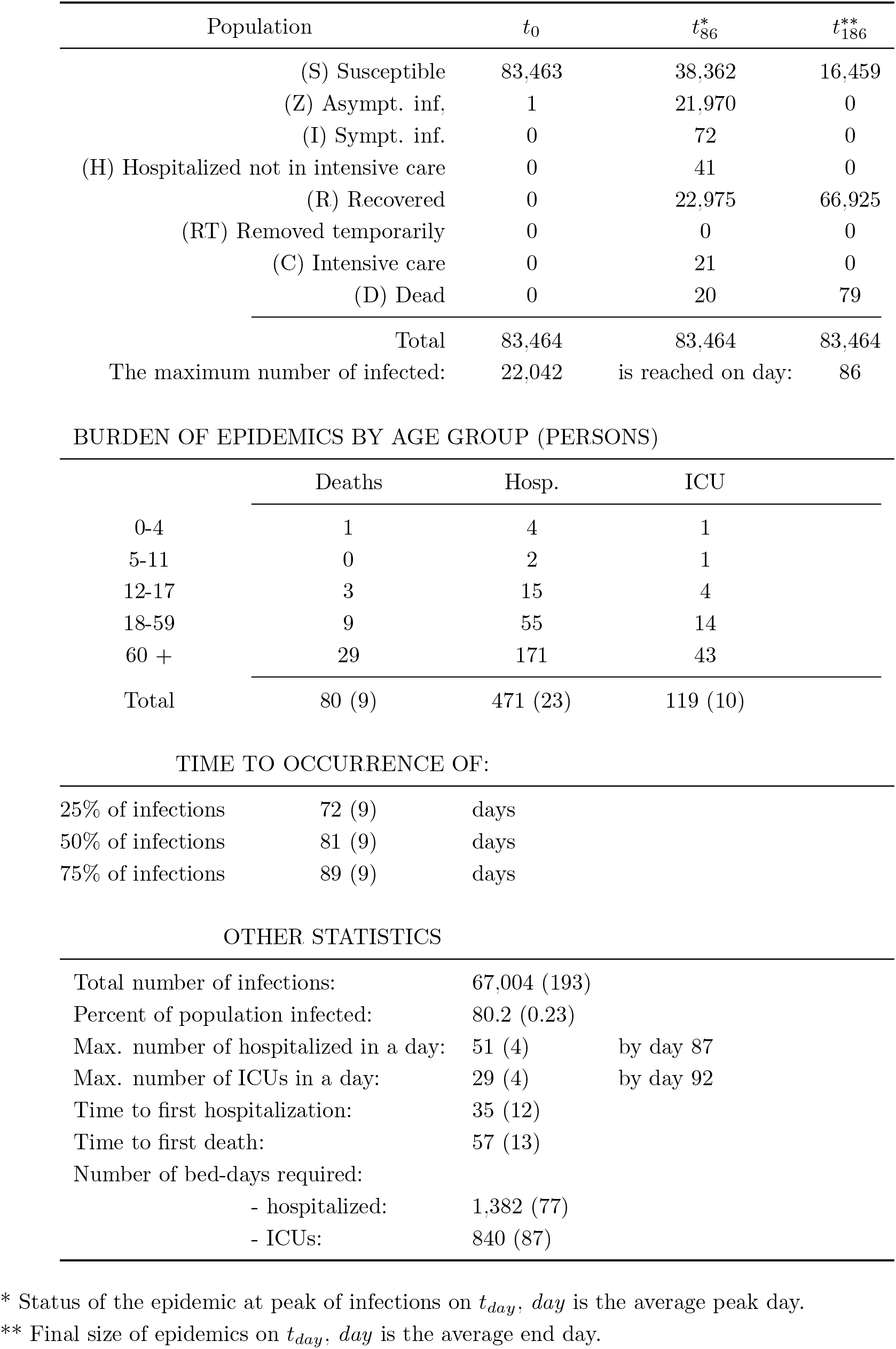
SIZ Model: *R*_0_ = 3, *θ* = 0, *C*_1_ = 0.33, *C*_2_ = 0.0, *C*_3_ = 3 (Numbers in parenthesis are SD)

| Population | $t_0$ | $t_{86}^*$ | $t_{186}^{**}$ |
| --- | --- | --- | --- |
| (S) Susceptible | 83,463 | 38,362 | 16,459 |
| (Z) Asympt. inf, | 1 | 21,970 | 0 |
| (I) Sympt. inf. | 0 | 72 | 0 |
| (H) Hospitalized not in intensive care | 0 | 41 | 0 |
| (R) Recovered | 0 | 22,975 | 66,925 |
| (RT) Removed temporarily | 0 | 0 | 0 |
| (C) Intensive care | 0 | 21 | 0 |
| (D) Dead | 0 | 20 | 79 |
| Total | 83,464 | 83,464 | 83,464 |
| The maximum number of infected: | 22,042 | is reached on day: | 86 |

BURDEN OF EPIDEMICS BY AGE GROUP (PERSONS)
|  | Deaths | Hosp. | ICU |
| --- | --- | --- | --- |
| 0-4 | 1 | 4 | 1 |
| 5-11 | 0 | 2 | 1 |
| 12-17 | 3 | 15 | 4 |
| 18-59 | 9 | 55 | 14 |
| 60 + | 29 | 171 | 43 |
| Total | 80 (9) | 471 (23) | 119 (10) |

TIME TO OCCURRENCE OF:
|  |  |  |
| --- | --- | --- |
| 25% of infections | 72 (9) | days |
| 50% of infections | 81 (9) | days |
| 75% of infections | 89 (9) | days |

OTHER STATISTICS
|  |  |  |
| --- | --- | --- |
| Total number of infections: | 67,004 (193) |  |
| Percent of population infected: | 80.2 (0.23) |  |
| Max. number of hospitalized in a day: | 51 (4) | by day 87 |
| Max. number of ICUs in a day: | 29 (4) | by day 92 |
| Time to first hospitalization: | 35 (12) |  |
| Time to first death: | 57 (13) |  |
| Number of bed-days required: |  |  |
| - hospitalized: | 1,382 (77) |  |
| - ICUs: | 840 (87) |  |

\* Status of the epidemic at peak of infections on $t_{day}$ , $day$ is the average peak day.\*\* Final size of epidemics on $t_{day}$ , $day$ is the average end day.

**Figure 5:**
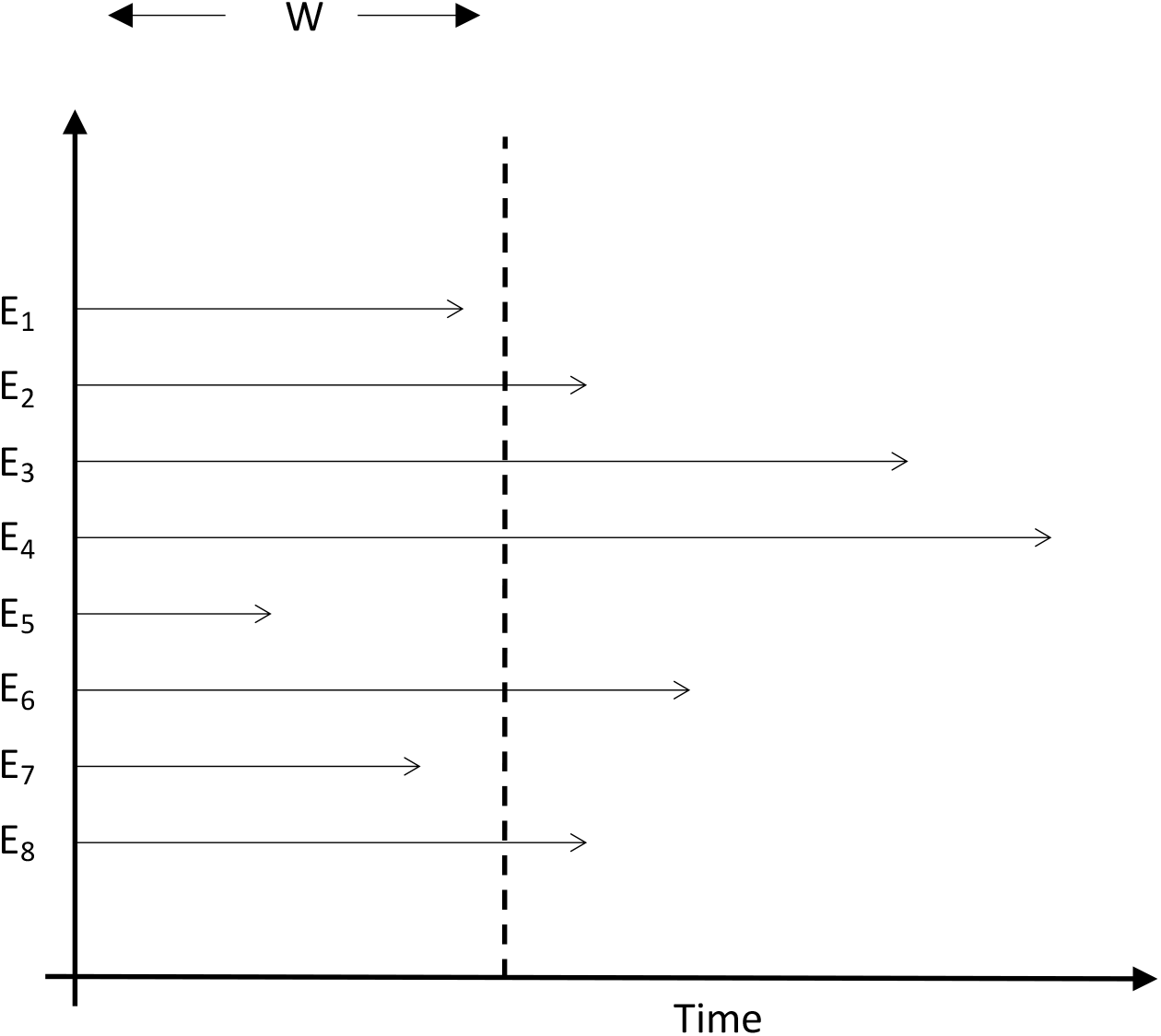
If inter-event times are exponential with parameter *µ*, the number of events in a window of size *W* has a Poisson distribution with parameter *Wµ*.

**Figure 6:**
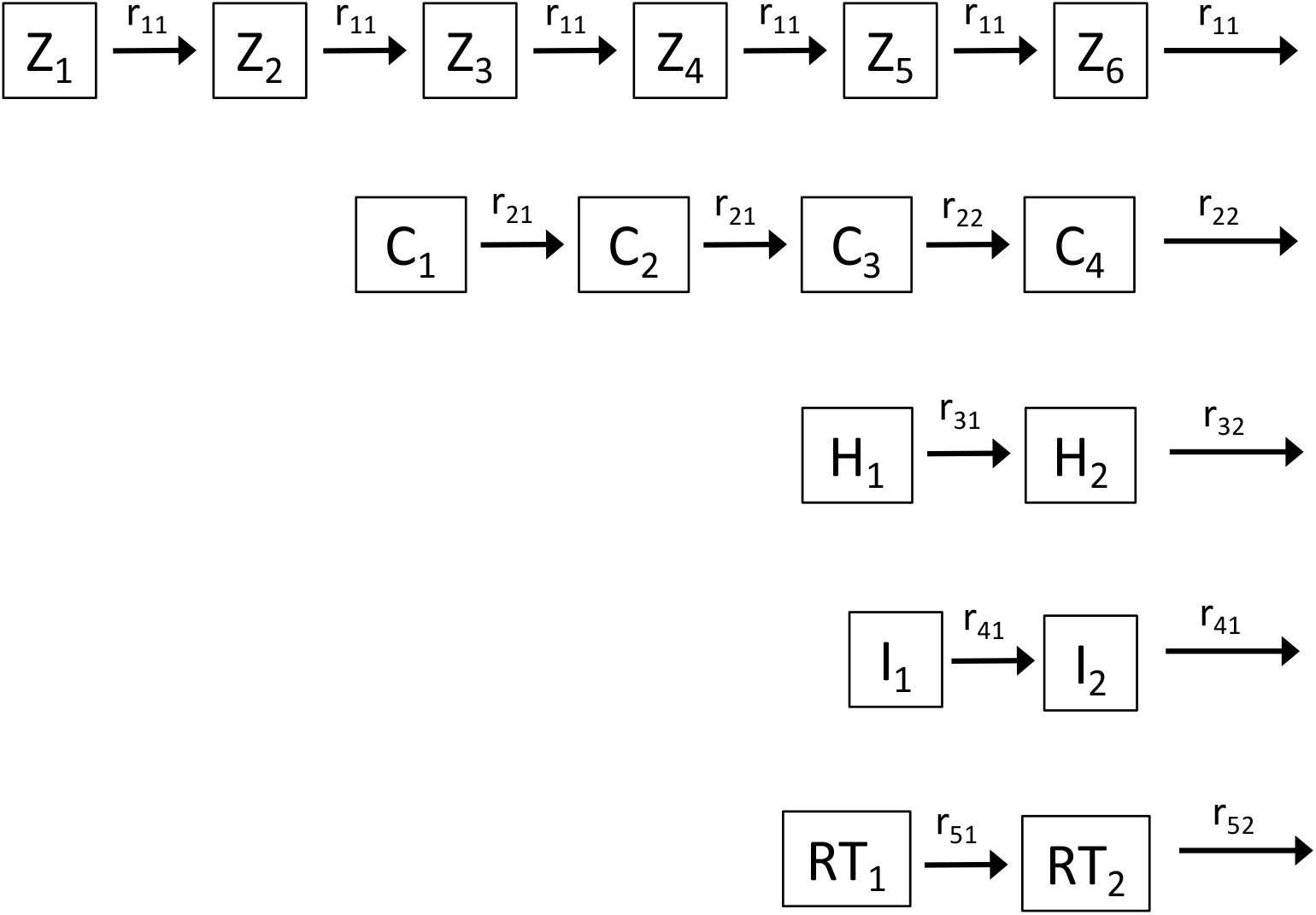
Division of stages in compartments to match the first two central moments. When an individual enters a stage, this person actually enters the first compartment of that stage and crosses all of the compartments and the indicated rates. This will cause the mean and variance of the time to cross all the compartments to be equal to the known mean and variance.

**Figure 7:**
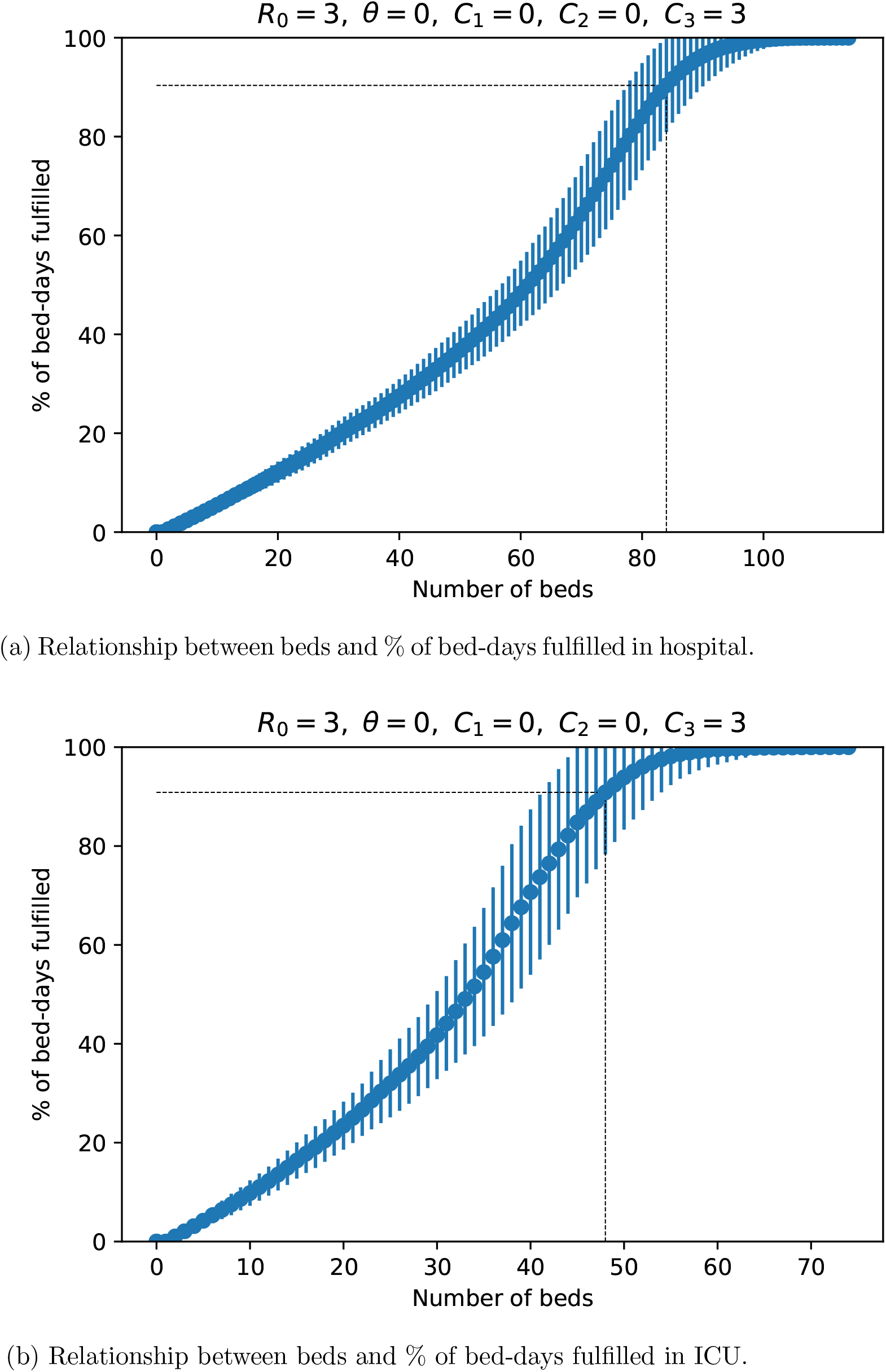
Number of beds required to fulfill a specific demand of bed-days. The dotted line points the required beds to achieve 90 % demand. The error bars correspond to standard deviations.

**Figure 8:**
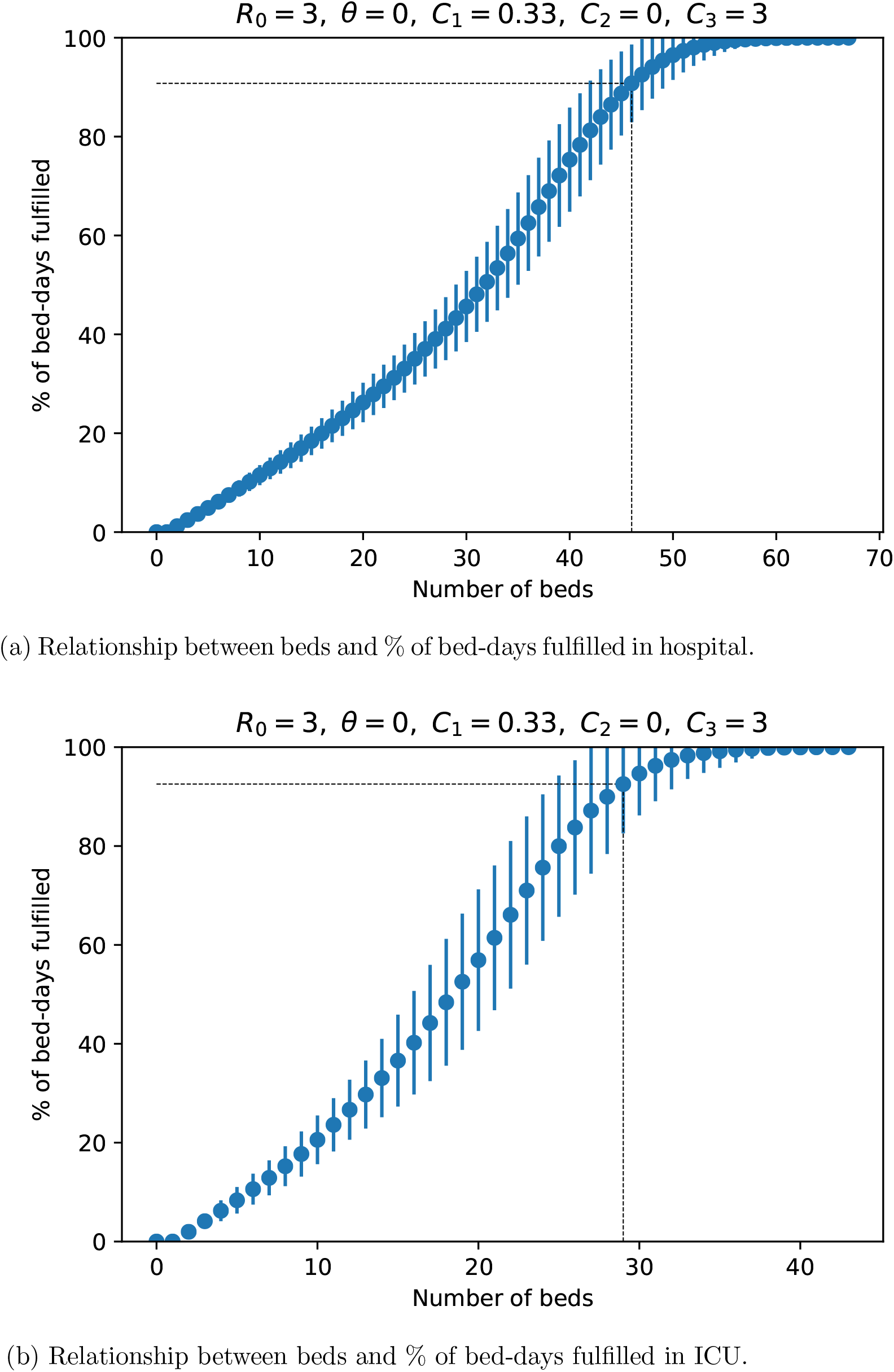
Number of beds required to fulfill a specific demand of bed-days. The dotted line points the required beds to achieve 90 % demand. The error bars correspond to standard deviations.

**Table 19:**
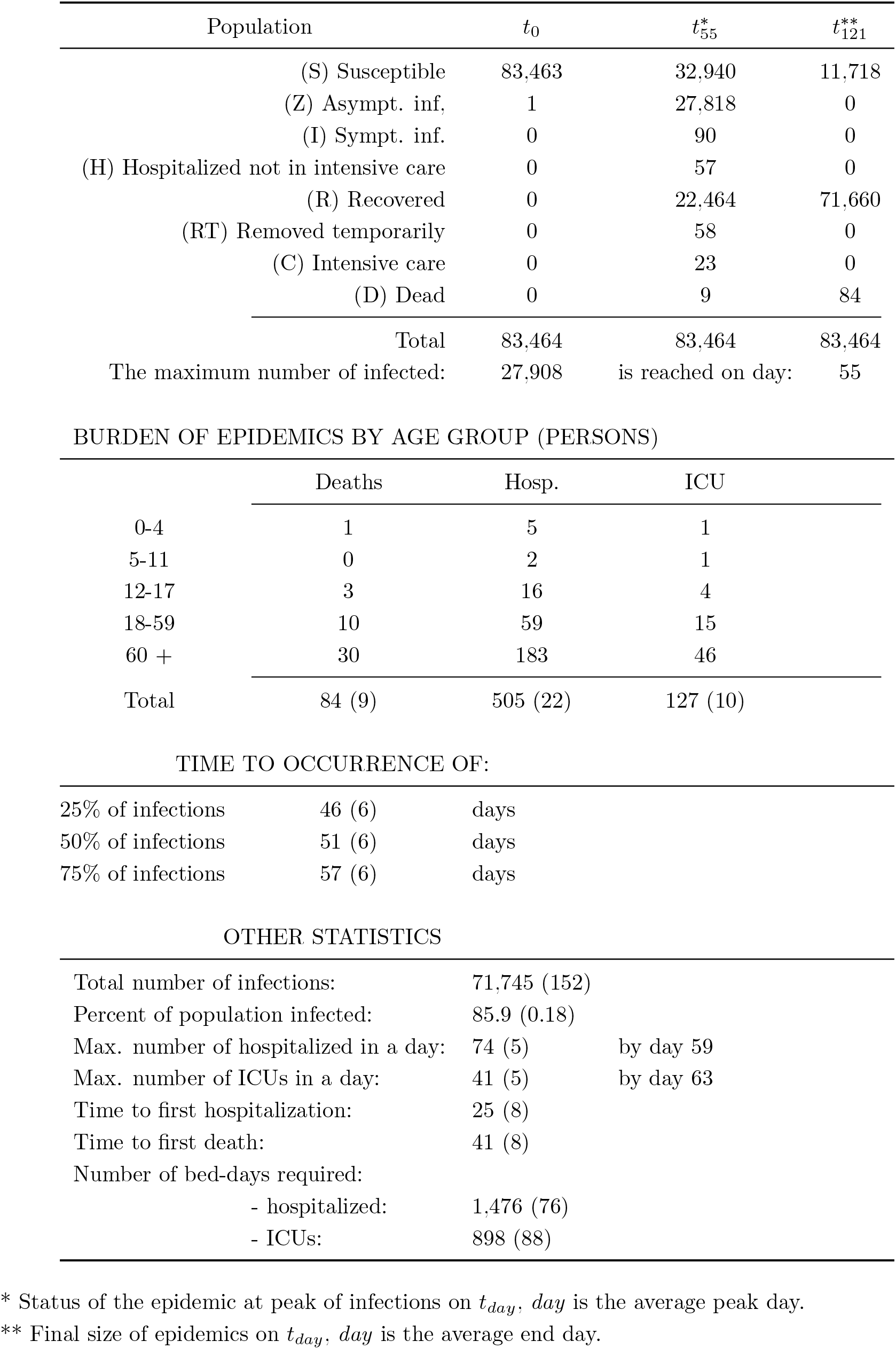
SIZ Model: *R*_0_ = 3, *θ* = 0.33, *C*_1_ = 0.0, *C*_2_ = 0.0, *C*_3_ = 3 (Numbers in parenthesis are SD)

| Population | $t_0$ | $t_{55}^*$ | $t_{121}^{**}$ |
| --- | --- | --- | --- |
| (S) Susceptible | 83,463 | 32,940 | 11,718 |
| (Z) Asympt. inf, | 1 | 27,818 | 0 |
| (I) Sympt. inf. | 0 | 90 | 0 |
| (H) Hospitalized not in intensive care | 0 | 57 | 0 |
| (R) Recovered | 0 | 22,464 | 71,660 |
| (RT) Removed temporarily | 0 | 58 | 0 |
| (C) Intensive care | 0 | 23 | 0 |
| (D) Dead | 0 | 9 | 84 |
| Total | 83,464 | 83,464 | 83,464 |
| The maximum number of infected: | 27,908 | is reached on day: | 55 |

BURDEN OF EPIDEMICS BY AGE GROUP (PERSONS)
|  | Deaths | Hosp. | ICU |
| --- | --- | --- | --- |
| 0-4 | 1 | 5 | 1 |
| 5-11 | 0 | 2 | 1 |
| 12-17 | 3 | 16 | 4 |
| 18-59 | 10 | 59 | 15 |
| 60 + | 30 | 183 | 46 |
| Total | 84 (9) | 505 (22) | 127 (10) |

TIME TO OCCURRENCE OF:
|  |  |  |
| --- | --- | --- |
| 25% of infections | 46 (6) | days |
| 50% of infections | 51 (6) | days |
| 75% of infections | 57 (6) | days |

OTHER STATISTICS
|  |  |  |
| --- | --- | --- |
| Total number of infections: | 71,745 (152) |  |
| Percent of population infected: | 85.9 (0.18) |  |
| Max. number of hospitalized in a day: | 74 (5) | by day 59 |
| Max. number of ICUs in a day: | 41 (5) | by day 63 |
| Time to first hospitalization: | 25 (8) |  |
| Time to first death: | 41 (8) |  |
| Number of bed-days required: |  |  |
| - hospitalized: | 1,476 (76) |  |
| - ICUs: | 898 (88) |  |

\* Status of the epidemic at peak of infections on $t_{day}$ , $day$ is the average peak day.\*\* Final size of epidemics on $t_{day}$ , $day$ is the average end day.

**Figure 9:**
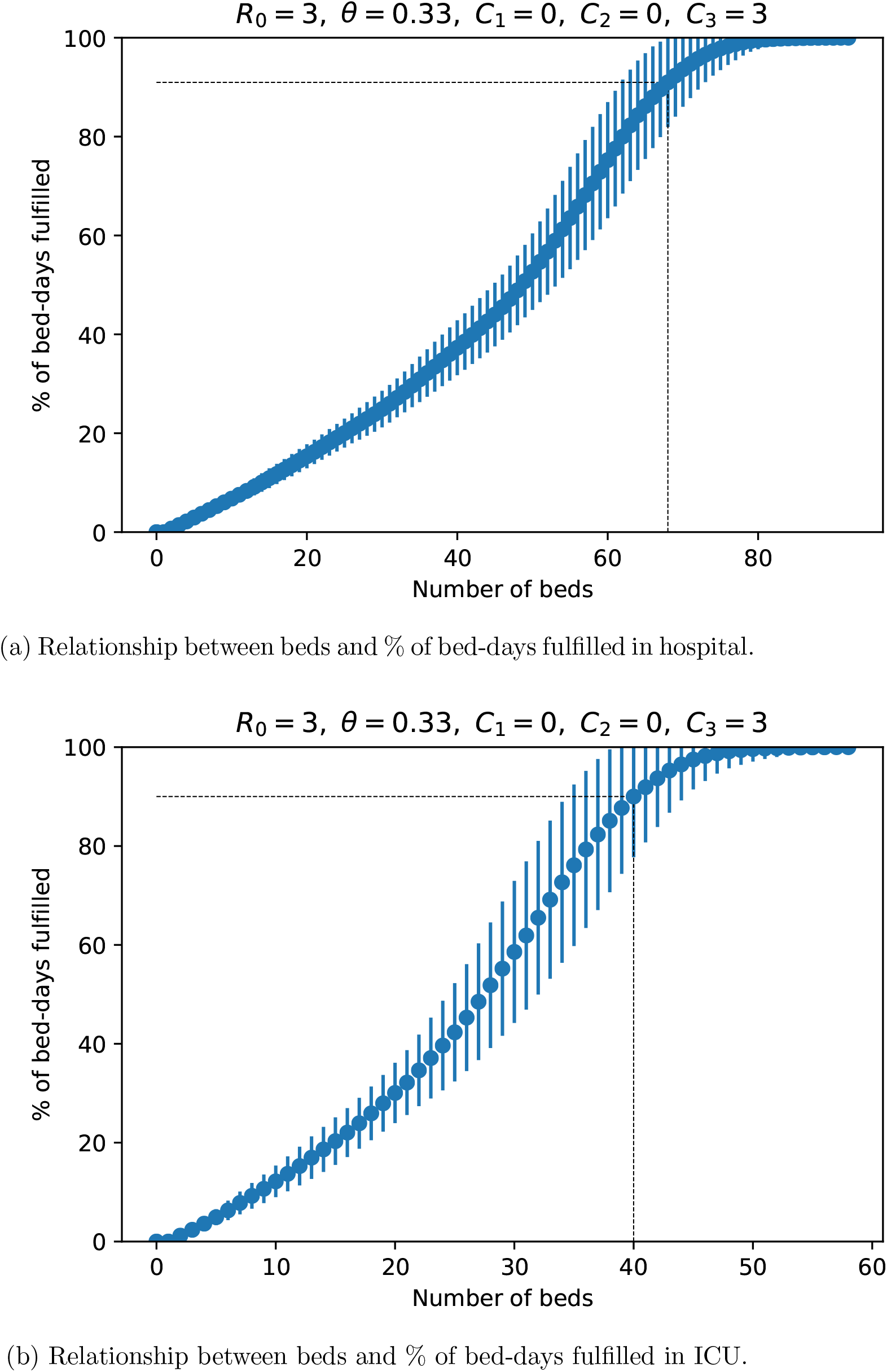
Number of beds required to fulfill a specific demand of bed-days. The dotted line points the required beds to achieve 90 % demand. The error bars correspond to standard deviations.

**Table 20:**
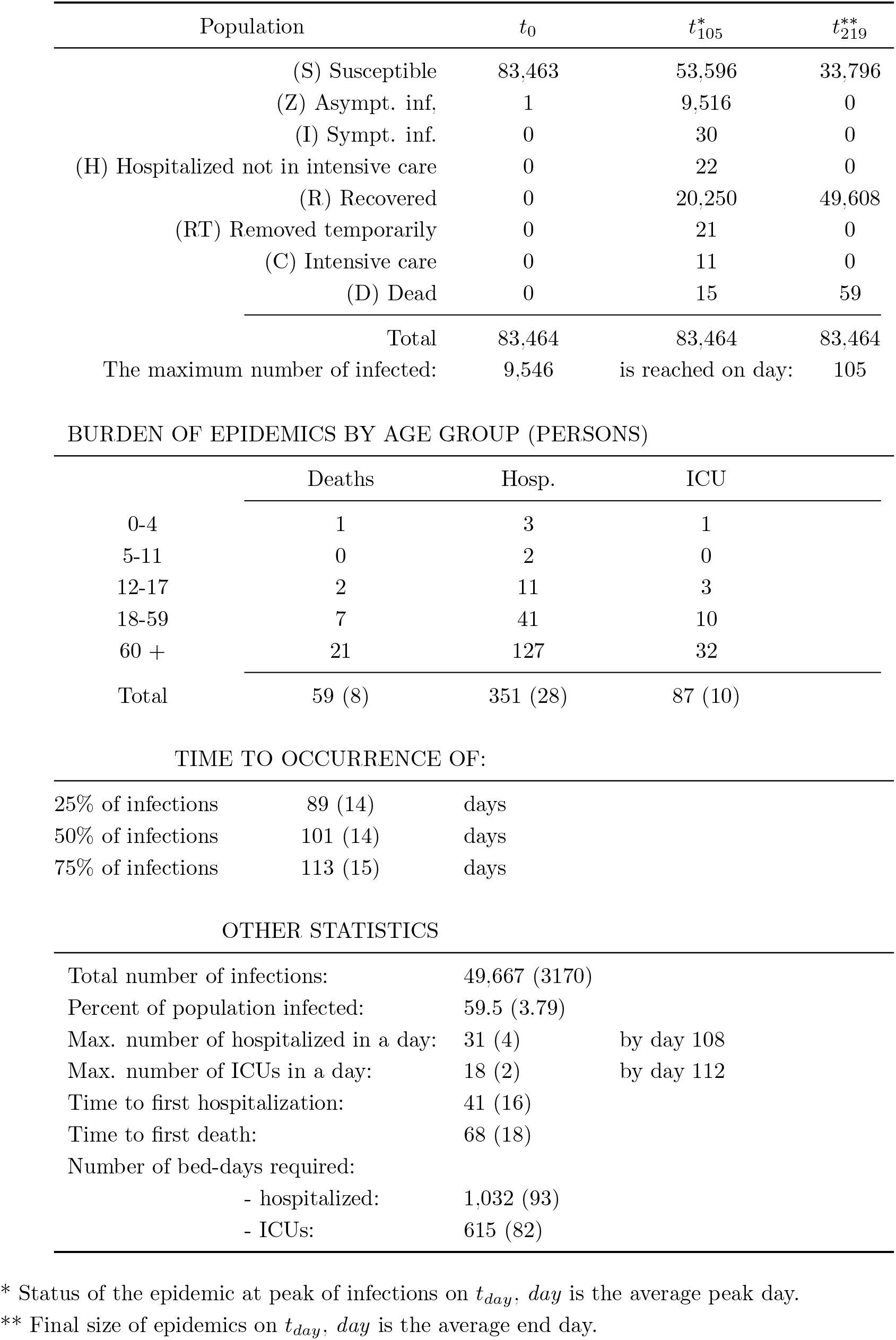
SIZ Model: *R*_0_ = 3, *θ* = 0.33, *C*_1_ = 0.33, *C*_2_ = 0.0, *C*_3_ = 3 (Numbers in parenthesis are SD)

| Population | $t_0$ | $t_{105}^*$ | $t_{219}^{**}$ |
| --- | --- | --- | --- |
| (S) Susceptible | 83,463 | 53,596 | 33,796 |
| (Z) Asympt. inf, | 1 | 9,516 | 0 |
| (I) Sympt. inf. | 0 | 30 | 0 |
| (H) Hospitalized not in intensive care | 0 | 22 | 0 |
| (R) Recovered | 0 | 20,250 | 49,608 |
| (RT) Removed temporarily | 0 | 21 | 0 |
| (C) Intensive care | 0 | 11 | 0 |
| (D) Dead | 0 | 15 | 59 |
| Total | 83,464 | 83,464 | 83,464 |
| The maximum number of infected: | 9,546 | is reached on day: | 105 |

BURDEN OF EPIDEMICS BY AGE GROUP (PERSONS)
|  | Deaths | Hosp. | ICU |
| --- | --- | --- | --- |
| 0-4 | 1 | 3 | 1 |
| 5-11 | 0 | 2 | 0 |
| 12-17 | 2 | 11 | 3 |
| 18-59 | 7 | 41 | 10 |
| 60 + | 21 | 127 | 32 |
| Total | 59 (8) | 351 (28) | 87 (10) |

TIME TO OCCURRENCE OF:
|  |  |  |
| --- | --- | --- |
| 25% of infections | 89 (14) | days |
| 50% of infections | 101 (14) | days |
| 75% of infections | 113 (15) | days |

OTHER STATISTICS
|  |  |  |
| --- | --- | --- |
| Total number of infections: | 49,667 (3170) |  |
| Percent of population infected: | 59.5 (3.79) |  |
| Max. number of hospitalized in a day: | 31 (4) | by day 108 |
| Max. number of ICUs in a day: | 18 (2) | by day 112 |
| Time to first hospitalization: | 41 (16) |  |
| Time to first death: | 68 (18) |  |
| Number of bed-days required: |  |  |
| - hospitalized: | 1,032 (93) |  |
| - ICUs: | 615 (82) |  |

\* Status of the epidemic at peak of infections on $t_{day}$ , $day$ is the average peak day.\*\* Final size of epidemics on $t_{day}$ , $day$ is the average end day.

**Figure 10:**
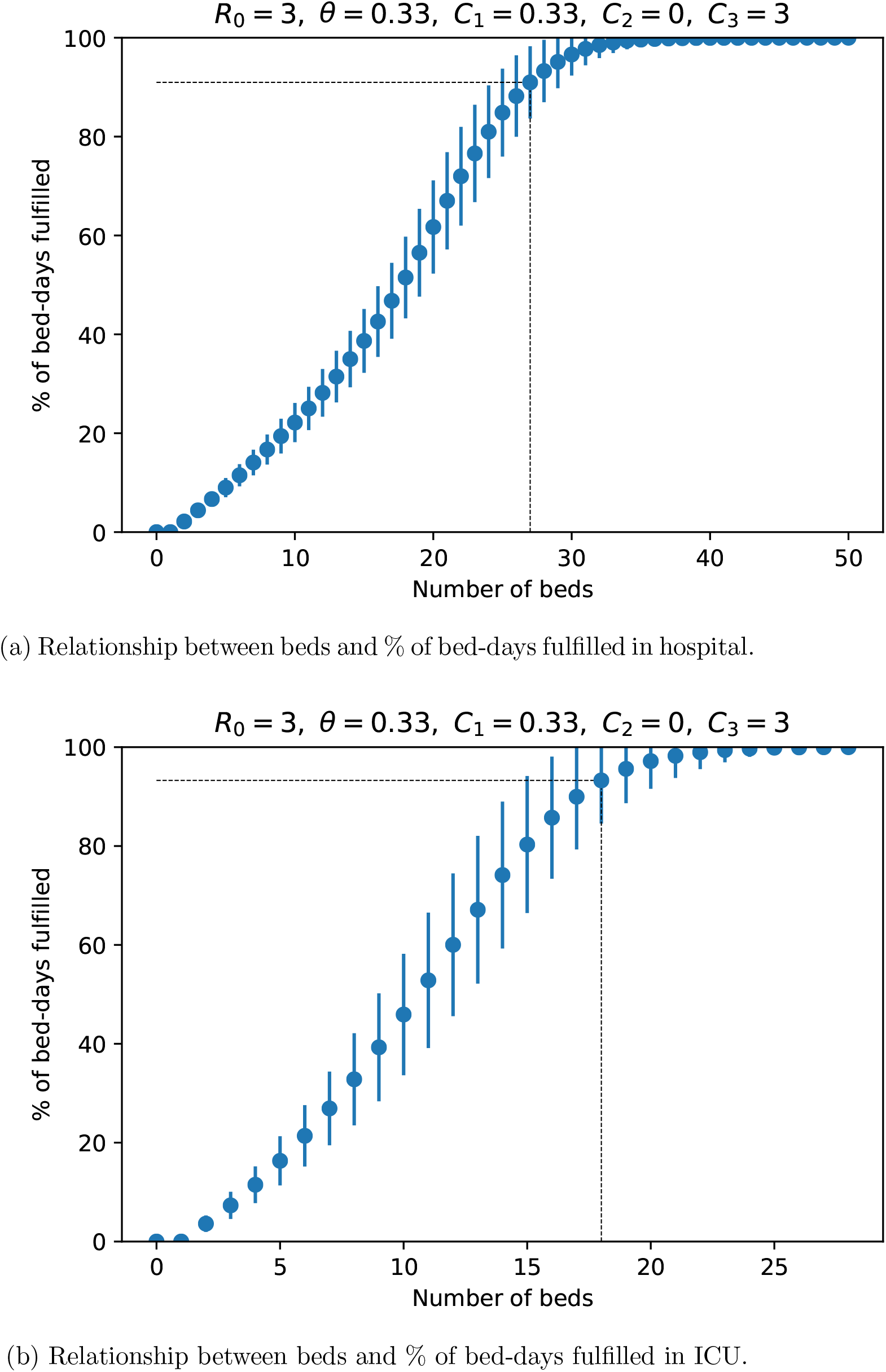
Number of beds required to fulfill a specific demand of bed-days. The dotted line points the required beds to achieve 90 % demand. The error bars correspond to standard deviations.

## Footnotes

1 A bed-day is a day during which a person is confined to a bed and in which the patient stays overnight in a hospital. (OECD Glossary of Statistical Terms https://stats.oecd.org/glossary/detail.asp?ID=194)

2 Note that the parameter dealing with reducing the contact-rate by self-awareness of infection had little effect on the outcome of the model and was excluded from the stochastic simulations.

3 https://data2.unhcr.org/en/documents/download/77747.

4 https://www.rescue.org/video/first-covid-19-case-confirmed-coxs-bazar-bangladesh

5 https://www.who.int/southeastasia/news/feature-stories/detail/who-partners-enhance-support-to-covid-19-response-in-rohingya-camps-in-coxs-bazar.

6 We used a database of suspected and confirmed COVID-19 cases maintained by the IMSS in Mexico (Instituto Mexicano de Seguro Social, or Mexican Institute of Social Security) that serves about 12 million persons, non-government workers, and cover roughly 10% of the Mexican population. The data on SARS-CoV-2 we used contains records of 26,039 confirmed cases from March 2, 2020 to May 25, 2020. We discarded all cases with symptoms not starting in the previous 14 days to the last reported case (May 25, 2020) and from these, included only cases with disease outcome as dead or recovered, leaving a database with 10,330 cases.

7 https://www.covid.is/data

8 shorturl.at/mR267

9 shorturl.at/mR267

## Notes

### Competing Interest Statement

The authors have declared no competing interest.

### Author Declarations

No IRB/oversight approval required

## References

[1] Gaurav Aggarwal, Isaac Cheruiyot, Saurabh Aggarwal, Johnny Wong, Giuseppe Lippi, Carl J Lavie, Brandon M Henry, and Fabian Sanchis-Gomar. Association of cardiovascular disease with coronavirus disease 2019 (covid-19) severity: A meta-analysis. Current Problems in Cardiology, page 100617, 2020.

[2] Jantien A Backer, Don Klinkenberg, and Jacco Wallinga. Incubation period of 2019 novel coronavirus (2019-ncov) infections among travellers from wuhan, china, 20–28 january 2020. Eurosurveillance, 25(5):2000062, 2020.

[3] Andrea Bobbio, András Horváth, and Miklós Telek. Matching three moments with minimal acyclic phase type distributions. Stochastic models, 21(2-3):303–326, 2005.

[4] David Champredon, Jonathan Dushoff, and David JD Earn. Equivalence of the erlang-distributed seir epidemic model and the renewal equation. SIAM Journal on Applied Mathematics, 78(6):3258–3278, 2018.

[5] Vincent CC Cheng, Shuk-Ching Wong, Vivien WM Chuang, Simon YC So, Jonathan HK Chen, Siddharth Sridhar, Kelvin KW To, Jasper FW Chan, Ivan FN Hung, Pak-Leung Ho, et al. The role of community-wide wearing of face mask for control of coronavirus disease 2019 (covid-19) epidemic due to sars-cov-2. Journal of Infection, 2020.

[6] Ensheng Dong, Hongru Du, and Lauren Gardner. An interactive web-based dashboard to track covid-19 in real time. The Lancet infectious diseases, 20(5):533–534, 2020.

[7] Steffen E Eikenberry, Marina Mancuso, Enahoro Iboi, Tin Phan, Keenan Eikenberry, Yang Kuang, Eric Kostelich, and Abba B Gumel. To mask or not to mask: Modeling the potential for face mask use by the general public to curtail the covid-19 pandemic. Infectious Disease Modelling, 2020.

[8] Christian Erikstrup, Christoffer Egeberg Hother, Ole Birger Vestager Pedersen, Kåre Mølbak, Robert Leo Skov, Dorte Kinggaard Holm, Susanne Sækmose, Anna Christine Nilsson, Patrick Terrence Brooks, Jens Kjaergaard Boldsen, Christina Mikkelsen, Mikkel Gybel-Brask, Erik Sørensen, Khoa Manh Dinh, Susan Mikkelsen, Bjarne Kuno Møller, Thure Haunstrup, Lene Harritshøj, Bitten Aagaard Jensen, Henrik Hjalgrim, Søren Thue Lillevang, and Henrik Ullum. Estimation of sars-cov-2 infection fatality rate by real-time antibody screening of blood donors. medRxiv, 2020.

[9] Xi He, Eric HY Lau, Peng Wu, Xilong Deng, Jian Wang, Xinxin Hao, Yiu Chung Lau, Jessica Y Wong, Yujuan Guan, Xinghua Tan, et al. Temporal dynamics in viral shedding and transmissibility of covid-19. Nature medicine, pages 1–4, 2020.

[10] Carlos Hernandez-Suarez and Efren Murillo-Zamora. Statistics associated with the lethality of covid-19 by age group and gender in mexico. medRxiv, 2020.

[11] Xuan Jiang, Simon Rayner, and Min-Hua Luo. Does sars-cov-2 has a longer incubation period than sars and mers? Journal of medical virology, 2020.

[12] Joseph T F Lau, Hiyi Tsui, Mason Lau, and Xilin Yang. Sars transmission, risk factors, and prevention in hong kong. Emerging infectious diseases, 10(4):587–592, 04 2004.

[13] Stephen A Lauer, Kyra H Grantz, Qifang Bi, Forrest K Jones, Qulu Zheng, Hannah R Meredith, Andrew S Azman, Nicholas G Reich, and Justin Lessler. The incubation period of coronavirus disease 2019 (covid-19) from publicly reported confirmed cases: estimation and application. Annals of internal medicine, 2020.

[14] Bruce G Lindsay, Ramani S Pilla, and Prasanta Basak. Moment-based approximations of distributions using mixtures: Theory and applications. Annals of the Institute of Statistical Mathematics, 52(2):215–230, 2000.

[15] Natalie M Linton, Tetsuro Kobayashi, Yichi Yang, Katsuma Hayashi, Andrei R Akhmetzhanov, Sung-mok Jung, Baoyin Yuan, Ryo Kinoshita, and Hiroshi Nishiura. Incubation period and other epidemiological characteristics of 2019 novel coronavirus infections with right truncation: a statistical analysis of publicly available case data. Journal of clinical medicine, 9(2):538, 2020.

[16] Hiroyuki Okamura and Tadashi Dohi. Ph fitting algorithm and its application to reliability engineering. Journal of the Operations Research Society of Japan, 59(1):72–109, 2016.

[17] Takayuki Osogami and Mor Harchol-Balter. Closed form solutions for mapping general distributions to quasi-minimal ph distributions. Performance Evaluation, 63(6):524–552, 2006.

[18] Raymond Pranata, Ian Huang, Michael Anthonius Lim, Eka Julianta Wahjoepramono, and Julius July. Impact of cerebrovascular and cardiovascular diseases on mortality and severity of covid-19–systematic review, metaanalysis, and meta-regression. Journal of Stroke and Cerebrovascular Diseases, page 104949, 2020.

[19] Yun Qiu, Xi Chen, and Wei Shi. Impacts of social and economic factors on the transmission of coronavirus disease 2019 (covid-19) in china. Journal of Population Economics, page 1, 2020.

[20] Camilla Rothe, Mirjam Schunk, Peter Sothmann, Gisela Bretzel, Guenter Froeschl, Claudia Wallrauch, Thorbjörn Zimmer, Verena Thiel, Christian Janke, Wolfgang Guggemos, et al. Transmission of 2019-ncov infection from an asymptomatic contact in germany. New England Journal of Medicine, 382(10):970–971, 2020.

[21] Hendrik Streeck, Bianca Schulte, Beate Kuemmerer, Enrico Richter, Tobias Hoeller, Christine Fuhrmann, Eva Bartok, Ramona Dolscheid, Moritz Berger, Lukas Wessendorf, Monika Eschbach-Bludau, Angelika Kellings, Astrid Schwaiger, Martin Coenen, Per Hoffmann, Markus Noethen, Anna-Maria Eis- Huebinger, Martin Exner, Ricarda Schmithausen, Matthias Schmid, and Beate Kuemmerer. Infection fatality rate of sars-cov-2 infection in a german community with a super-spreading event. medRxiv, 2020.

[22] Giovanni Targher, Alessandro Mantovani, Xiao-Bo Wang, Hua-Dong Yan, Qing-Feng Sun, Ke-Hua Pan, Christopher D Byrne, Kenneth I Zheng, Yong-Ping Chen, Mohammed Eslam, et al. Patients with diabetes are at higher risk for severe illness from covid-19. Diabetes & Metabolism, 2020.

[23] Shaun Truelove, Orit Abrahim, Chiara Altare, Stephen A. Lauer, Krya H. Grantz, Andrew S. Azman, and Paul Spiegel. The potential impact of covid-19 in refugee camps in bangladesh and beyond: A modeling study. PLOS Medicine, 17(6):1–15, 06 2020.

[24] M Villa, JF Myers, and F Turkheimer. Covid-19: Recovering estimates of the infected fatality rate during an ongoing pandemic through partial data. medrxiv. 2020.

[25] Fei Zhou, Ting Yu, Ronghui Du, Guohui Fan, Ying Liu, Zhibo Liu, Jie Xiang, Yeming Wang, Bin Song, Xiaoying Gu, et al. Clinical course and risk factors for mortality of adult inpatients with covid-19 in wuhan, china: a retrospective cohort study. The lancet, 2020.

